# Household contact management of drug-resistant tuberculosis: a global cost-effectiveness modelling analysis of expanded preventive treatment by age

**DOI:** 10.64898/2026.10.01.26364489

**Authors:** Nyashadzaishe Mafirakureva, Christopher F McQuaid, Martin Harker, Anthony Garcia-Prats, Anneke C Hesseling, James A Seddon, Peter J Dodd

## Abstract

**Background:** Tuberculosis preventive treatment (TPT) with levofloxacin reduces tuberculosis disease incidence in multidrug-resistant or rifampicin-resistant (MDR/RR)-tuberculosis household contacts. However, with age, disease progression risk falls and the frequency of adverse events (AE) rises.

**Methods:** To inform decision-makers, we evaluated the cost-effectiveness of expanding the use of MDR/RR-TPT by age, with and without tests of tuberculosis infection. We modelled MDR/RR-household contact management (HHCM) in 189 countries, incorporating age-dependent risks of disease progression, death and AEs. Incremental cost-effectiveness ratios were calculated as USD/disability-adjusted life year (DALY) averted, and as a fraction of per capita gross domestic product (GDP).

**Findings:** Compared to baseline of MDR/RR-HHCM with TPT for those <15 years, adding TPT up to 45 years was cost-effective at a threshold of 30% GDP in 160/189 modelled countries including 17 of the 30 highest incidence countries. Of these 160 countries, the intervention was cost-saving in 46. Strategies targeting TPT to those testing positive for *M. tuberculosis* infection were dominated by those without testing. Globally, full use of MDR/RR-HHCM with TPT for all would avert 11,900 (10,400 to 13,800) episodes of tuberculosis, 9,960 (9,100 to 10,900) deaths, and 289,000 (262,000 to 321,000) DALYs, but would result in 71,000 (68,000 to 74,000) SAEs and cost $359 million (294 to 449).

**Interpretation:** Expanding MDR/RR-TPT use to older ages without testing appears cost-effective in most settings, but may require substantial resources and result in large numbers of AEs.

**Funding:** Unitaid

## Background

An estimated 10.7 million individuals developed tuberculosis in 2024, of whom 390,000 had multidrug-resistant (MDR) or rifampicin-resistant (RR)-tuberculosis.^1^ To substantially reduce the burden of MDR/RR-TB, combinations of complementary approaches will be required. First, new approaches are needed to identify individuals with tuberculosis who have not presented to health services and remain undiagnosed. Second, increased use of preventive strategies is needed, including vaccines and tuberculosis preventive treatment (TPT). Household contact management (HHCM) provides an opportunity to conduct active case-finding (ACF) of household members who have already developed tuberculosis, as well as provide TPT to contacts at high risk of future disease progression. Currently, the World Health Organization (WHO) and most national tuberculosis guidelines advise the evaluation of household contacts of individuals with newly diagnosed, infectious tuberculosis, in order to identify prevalent tuberculosis as well as provision of TPT. However, throughout much of the world, this is poorly implemented for household contacts of both drug-susceptible and MDR/RR-TB.

The provision of TPT to reduce the risk of progression from tuberculosis infection to disease has been carried out for over 60 years and good evidence exists for the efficacy and safety of regimens to prevent disease in those exposed to drug-susceptible tuberculosis.^2^ Ascertainment of tuberculosis infection, through tuberculin skin testing (TST) or interferon-gamma release assays (IGRAs), is rarely carried out in high tuberculosis-burden settings, and provision of TPT is commonly provided to those deemed to have sustained and significant exposure, considered an acceptable proxy for infection. However, until recently there were limited data to support the use of TPT for individuals exposed to MDR/RR-tuberculosis. Given that 147,592 individuals were diagnosed with MDR/RR-tuberculosis in 2024,^1^ the development of TPT regimens for those exposed to MDR/RR-tuberculosis remains a pressing priority.

In 2024, results from the TB-CHAMP^3^ and VQUIN^4^ trials were published. TB-CHAMP recruited children <5 years, and those 5 to <18 years with a positive IGRA or living with HIV, who had been living in the household of an individual with MDR-tuberculosis, and randomised them to receive levofloxacin or placebo daily for 6 months. VQUIN MDR recruited mainly IGRA-positive adults who had household exposure to MDR-tuberculosis and also randomised them to levofloxacin or placebo daily for 6 months. Integrated results from the two trials demonstrated a significant reduction in the risk of incident tuberculosis in contacts receiving levofloxacin.^5^ These results were considered by the WHO and incorporated into global guidance. The current WHO guidance states that children <5 years who are exposed to tuberculosis in their household should be given TPT, while individuals ≥5 years could be given TPT.^2^ The guidance suggests that tests of tuberculosis infection can be used for TPT decision-making, and that levofloxacin TPT should be used for household contacts of MDR/RR-tuberculosis.

These recommendations have been developed to cover settings with very different tuberculosis epidemiologies and resources. However, challenges remain for national Departments of Health or tuberculosis programmes, in interpreting guidance statements and deciding how best to apply them in their context. To plan how to conduct HHCM, countries need to understand the relative impact of screening for prevalent disease versus TPT, which contacts to prioritise for TPT, and whether tests of tuberculosis infection should be used. For countries with limited budgets, it is critical to quantify the impact, adverse effects, costs and cost-effectiveness of different approaches.

Age is a central consideration in TPT policy design. Age influences the risk of progression to disease following infection, the rate of adverse events due to TPT, and the case fatality rate from treated or untreated tuberculosis. In addition, a positive test of tuberculosis infection only indicates infection at some point during the lifetime of the individual. As recent exposure is more likely to lead to incident tuberculosis, it is important to understand the way age influences how likely a positive test of infection is to be due to recent infection. Given the global heterogeneity in country demographies, tuberculosis and MDR-tuberculosis epidemiologies, and costing parameters, it is likely that different countries will choose to implement the WHO guidance in different ways. We therefore aimed to develop a model to quantify the impact, cost and cost-effectiveness of different HHCM and TPT strategies to inform the implementation of these activities.

## Methods

### Interventions

We consider levofloxacin-based TPT for household contacts of MDR/RR-tuberculosis. A test for tuberculosis infection can be used to target TPT towards a smaller number of people with a higher risk of progression. For each age group (0-4, 5-14, 15-24, 25-34, 35-44, 45-54, 55-64 and 65+ years), we therefore consider four intervention conditions: 1) no HHCM; 2) HHCM without use of TPT (i.e. ACF only); 3) HHCM with TPT for PLHIV and those with positive TST; 4) HHCM with TPT for all. Within each age group, we consider an idealised intervention with perfect coverage and linkage to all aspects of care.

To inform expanded use of TPT in adult age groups, we consider the changes from sequentially offering TPT to new age groups, starting from a baseline of HHCM with TPT for those aged <15 years. This builds on previous work,^6,7^ and recognizes children as having high risks of progression and missed diagnosis. Second, we consider the changes compared to a baseline of no HHCM within each age group. These comparisons indicate total resource and health impacts of both the ACF and TPT components of HHCM, and how these vary by age.

Since countries are the relevant jurisdiction for policy decision-making, and because their costs, MDR/RR-tuberculosis epidemiology, fluoroquinolone resistance, and demography all vary, we calculate our outcomes on a country-by-country basis.

### Modelling approach

We build on previous modelling work in this area, combining models of household structure with data on tuberculosis diagnoses, estimates of tuberculosis drug-resistance patterns and concordance, and models of outcomes and costs across 189 countries in 2024 (see Figure 1).

**Figure 1:**
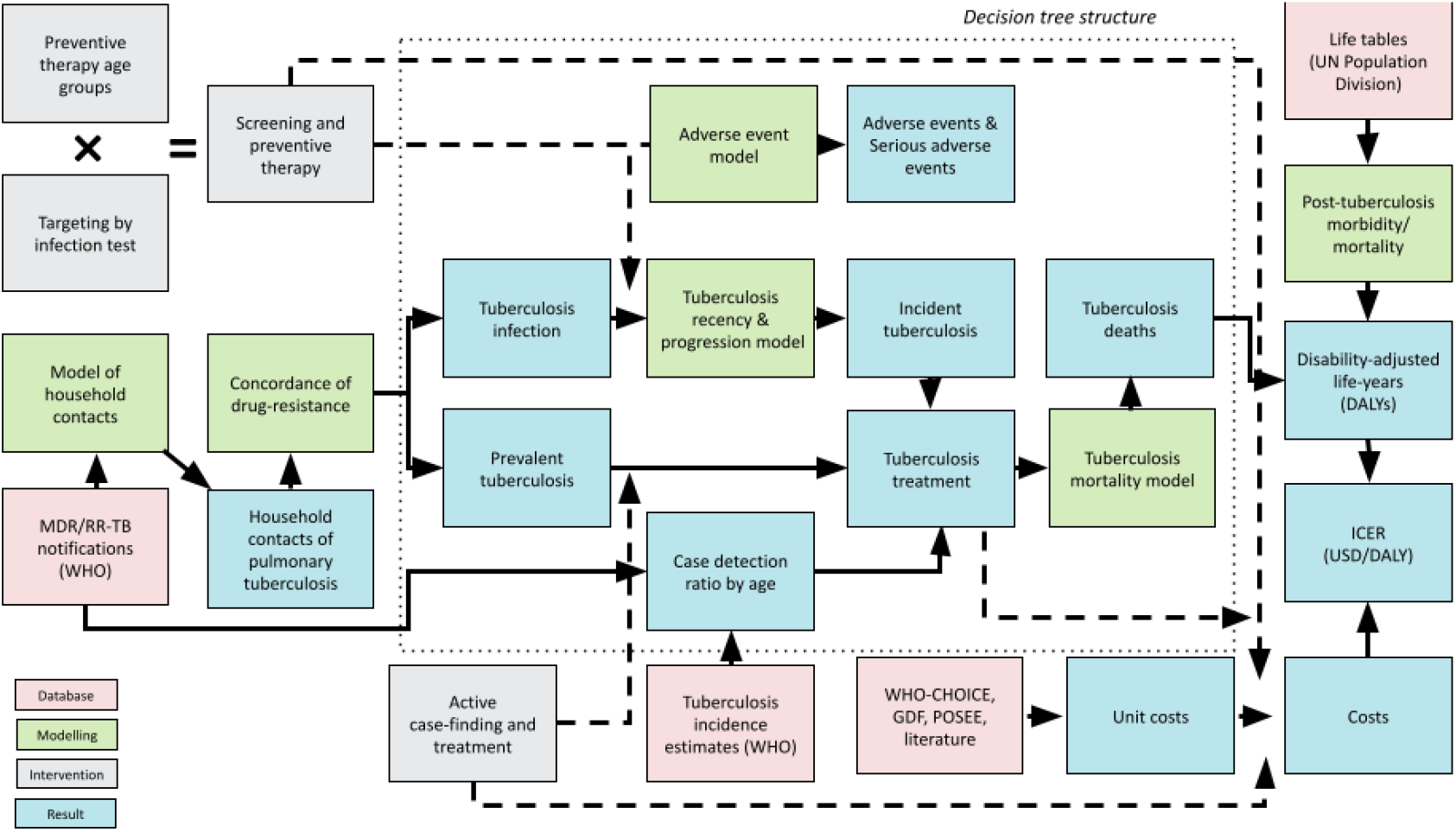
Overview of data sources and analysis pipeline. Analyses were performed at country level. Age dependence not shown. ICER = Incremental Cost-Effectiveness Ratio; WHO = World Health Organization; GDF = Global Drug Facility; POSEE = Pediatric TB Operational and Sustainability Expertise Exchange; WHO-CHOICE = WHO-CHOosing Interventions that are Cost-Effective

The household contact model was based on an updated multivariate regression analysis of 76 DHS data sets (see Appendix section 1.1.1). The resulting model allowed prediction in each country of the number of household members in each age group for an index individual of a given age and sex. This model was combined with age- and sex-stratified tuberculosis notification data in each country, and country estimates of the fraction of notified tuberculosis episodes that were identified as being MDR/RR-tuberculosis (see Appendix section 1.1.2). The result of this modelling stage was a population of household contacts on which various intervention options could act.

While a positive TST or IGRA in younger household contacts is most likely due to recent infection, older age groups in high tuberculosis prevalence settings are more likely to have a positive test from non-recent infection, representing a lower risk of progression to tuberculosis disease. To capture this effect, we introduced a novel analytical model of tuberculosis infection that calculates the probability a positive TST/IGRA in a household contact represents recent (re)infection (within 2 years) by age, accounting for partial protection against re-infection. This model was parametrized with country-specific background tuberculosis infection rates derived from Houben and Dodd,^8^ and household tuberculosis infection rates from Fox et al.,^9^ and was combined with rates of progression for recent and non-recent infection to model risks of tuberculosis disease incidence (see Appendix section 1.2).

To account for varying levels of fluoroquinolone-resistance by country and potential discordance in drug-resistance type between contacts and the index diagnosis, we followed an approach described previously.^6^ We estimated the fraction of MDR/RR-tuberculosis that is fluoroquinolone-resistant in each country using data reported to the WHO, and applied a probability that strains are concordant derived from Chiang et al et al.^10^ (see Appendix section 1.1.2).

The efficacy of TPT in preventing incident tuberculosis was based on a Bayesian analysis of the VQUIN MDR and TB-CHAMP trials,^5^ modelled as described in Seddon et al.^7^ The probability that prevalent or incident tuberculosis would be detected and appropriately treated followed a previously-developed approach based on WHO estimates that accounted for an increased probability of diagnosis in households with an index tuberculosis diagnosis.^6^ Antituberculosis treatment (ATT) success rates were based on WHO estimates, and treatment was assumed ineffective when not matched with drug-resistance type.

A number of features were modelled as depending on age, including the prevalence of tuberculosis infection and disease, as well as the risks of incident tuberculosis (see above and Appendix section 1.2). HIV-infection status in contacts is based on Martinez et al.^11^ for children and Calderwood et al.^12^ for adults (see Appendix section 1.1.2). We modelled risks of death with and without appropriate ATT as increasing with age, using data from Osman et al.^13,14^ (see Appendix section 1.4). Finally, we modelled increasing risks of adverse events and serious adverse events associated with TPT as increasing with age, based on data from Yuen et al.^15^ (see Appendix section 1.3).

### Health-economic approach

We evaluated the cost-effectiveness of sequentially offering TPT to additional age groups, beginning with TPT for only to those aged <15 years and ACF for all ages. HIV testing and TST costs were included for strategies targeting TPT in this way. TPT costs comprised drugs, follow-up visits, treatment monitoring, and adverse events management. Lastly, ATT costs included drugs, outpatient care, inpatient care and national tuberculosis programme (NTP) costs as reported by countries to WHO in 2024. ATT-related adverse event costs were not separately included and were assumed to be captured within NTP-reported ATT costs. Post-tuberculosis costs were excluded. All costs were calculated using publicly available information and adjusted for inflation to 2024 USD dollars. See Appendix section 2 for more details.

We also evaluated the cost-effectiveness of each intervention strategy compared to a baseline of no HHCM, including costs for household visits, contact screening and tuberculosis evaluation (outpatient visit, chest x-ray, Xpert MTB/RIF test for all symptomatic contacts).

All economic analyses were from a health system perspective. This analysis was not pre-specified in a health economic analysis plan.

Health outcomes include co-prevalent and incident tuberculosis, co-prevalent and incident tuberculosis-related deaths, discounted (at 3% per year) life-years lost and disability-adjusted life years (DALYs). Post-tuberculosis impacts on health-related quality of life and mortality are described in Appendix section 1.5.2.

### Metrics calculated

We used probabilistic sensitivity analysis (PSA) to include uncertainty in all results, with a sample of 1000 parameters for all country/intervention combinations. A decision tree model was used to calculate costs and outcomes across the PSA dataset, and means and 95% quantiles were reported as central and 95% uncertainty intervals (95%UIs). Analyses were run in R 4.6.0 and code and data to reproduce results are available on GitHub.

TPT expansion results reported the incremental and cumulative outputs from expanding TPT use to each adult age group in order of decreasing cost-effectiveness. Incremental cost-effectiveness ratios (ICERs) per DALY averted were reported as a percentage of per capita gross domestic product (GDP).

Relative to the baseline of no HHCM in each age group, we additionally calculated incremental resource use (household contacts screened; TPT courses; ATT courses; costs), and incremental health outcomes (tuberculosis incidence; tuberculosis deaths; adverse events and serious adverse events; DALYs).

## Findings

The probability a positive TST/IGRA represents recent (re)infection in a household contact is shown in Figure 2 for three different levels of household attack rate (quantified as the probability of household infection in contacts without previous infection). For ages under 15 years (x-axis) in low tuberculosis transmission settings, defined by tuberculosis infection prevalence below 10% at age 30 years (y-axis), over 87% of positive TST/IGRAs represent recent infection. This proportion declines with age and increasing levels of background transmission; in high tuberculosis transmission settings (infection prevalence over 40% at 30 years), a minority of positive tests in those over aged over 30 years represent recent infection (<40%).

**Figure 2:**
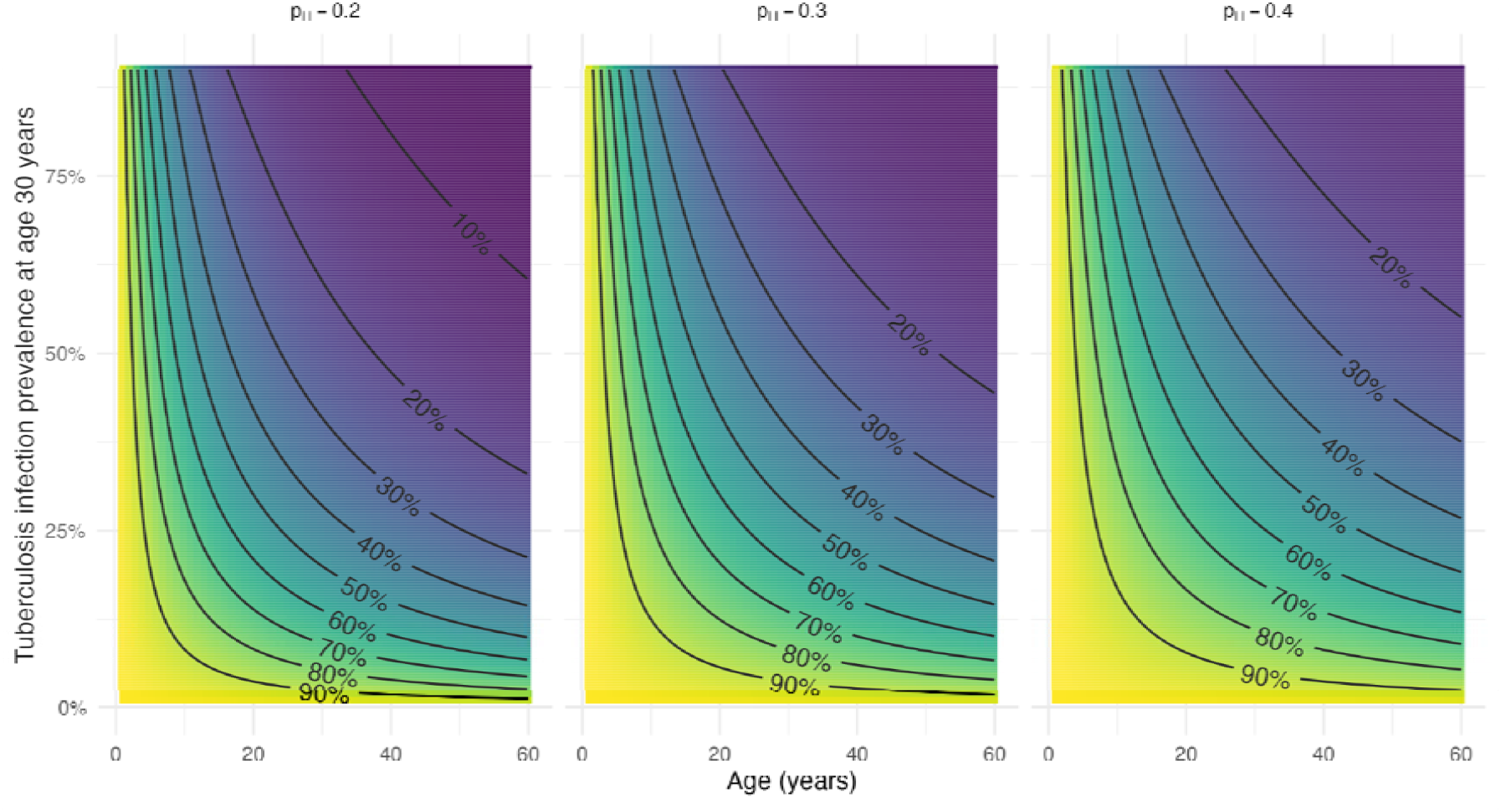
Percentage of positive tuberculosis infection tests in household contacts representing (re)infection within two years, and thus higher progression risk, as a function of age (x-axis) and community transmission (quantified as tuberculosis infection prevalence at 30 years; y-axis). The three panels represent different assumptions for the probability that an uninfected household contact would be infected by the index episode before household contact management (*p*_*H*_).

The globally-aggregated pathway for sequentially expanding TPT use through adult age groups in order of cost-effectiveness is shown in Figure 3. Globally, the optimal order for expanded TPT use corresponded with increasing age, with worse cost-effectiveness in older age groups. TPT without use of TST/IGRA dominated test-based targeting, conferring both larger health benefits and lower incremental costs. Appendix figure A5 shows this expansion pathway for 30 high tuberculosis incidence countries, including where expansion would stop being cost-effective at a cost-effectiveness threshold of 30% GDP. In 17/30 this meant TPT would be used up to the age of 45 years.

**Figure 3:**
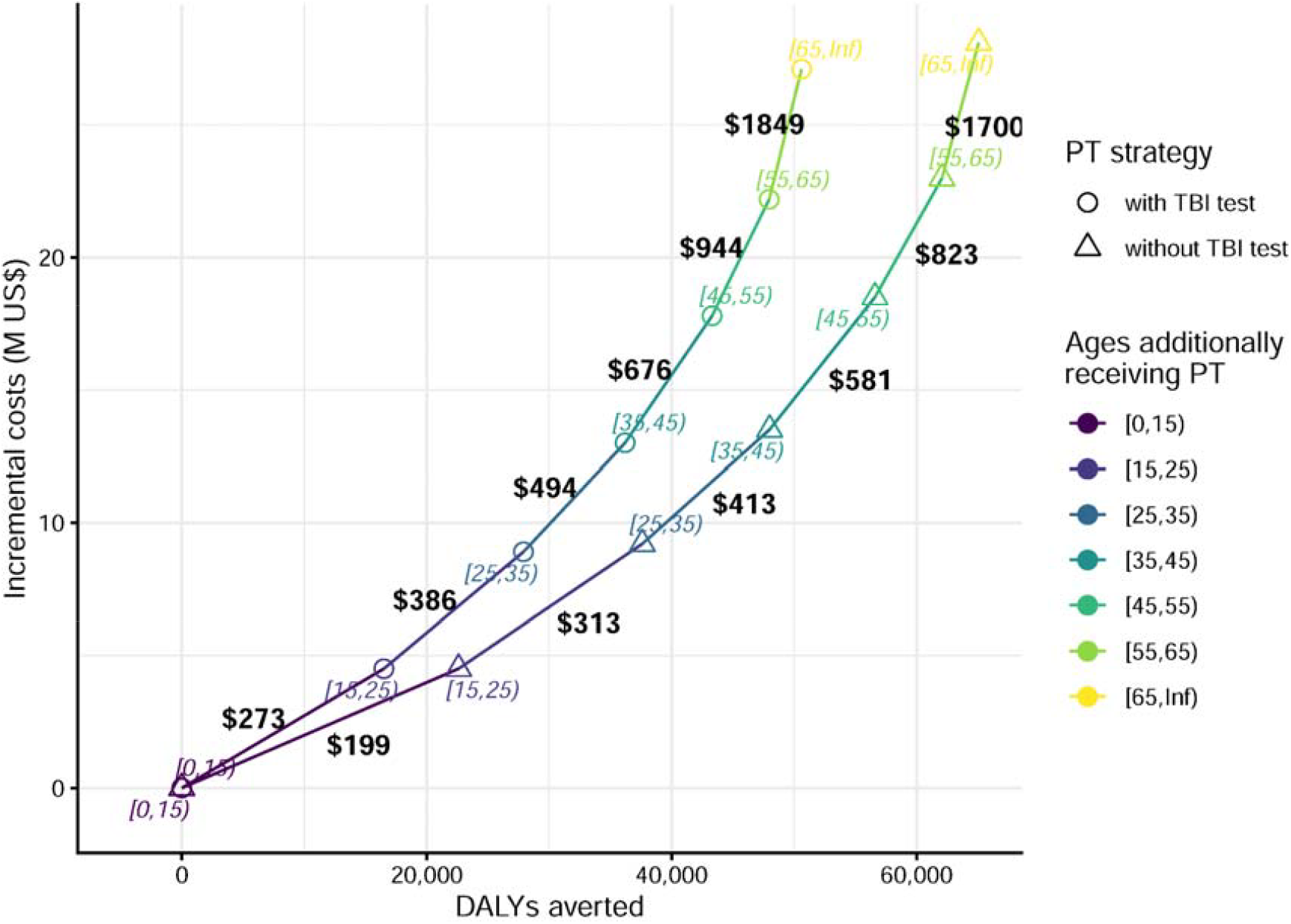
Global aggregate cost-effectiveness of expanding use of preventive treatment across contact age groups in households receiving multidrug-resistant/rifampicin-resistant tuberculosis contact management, with and without tuberculosis infection testing. Numbers on each line segment are the incremental cost-effectiveness ratio in US$ per disability-adjusted life-year (DALY) averted for expanding preventive treatment use to the most cost-effective choice of next age group. The origin represents active case-finding for all and preventive treatment for all <15 years. TBI = tuberculosis infection; PT = preventive treatment.

Figure 4 shows the ICERs of expanding TPT use up to the age of 45 years compared to a baseline of ACF for all and TPT for all aged <15 years, as a percentage of GDP in each country. For 46/189 countries, typically in the global north, the intervention is cost-saving. For 114/189 further countries the intervention is <30% of GDP, which, combined with countries where the intervention is cost-saving would comprise 84.6% of all countries, representing 75.6% of global tuberculosis incidence in 2024. A further 21/189 countries had ICERs below 50% GDP, with only 6/187 having ICERs exceeding 100% GDP.

**Figure 4:**
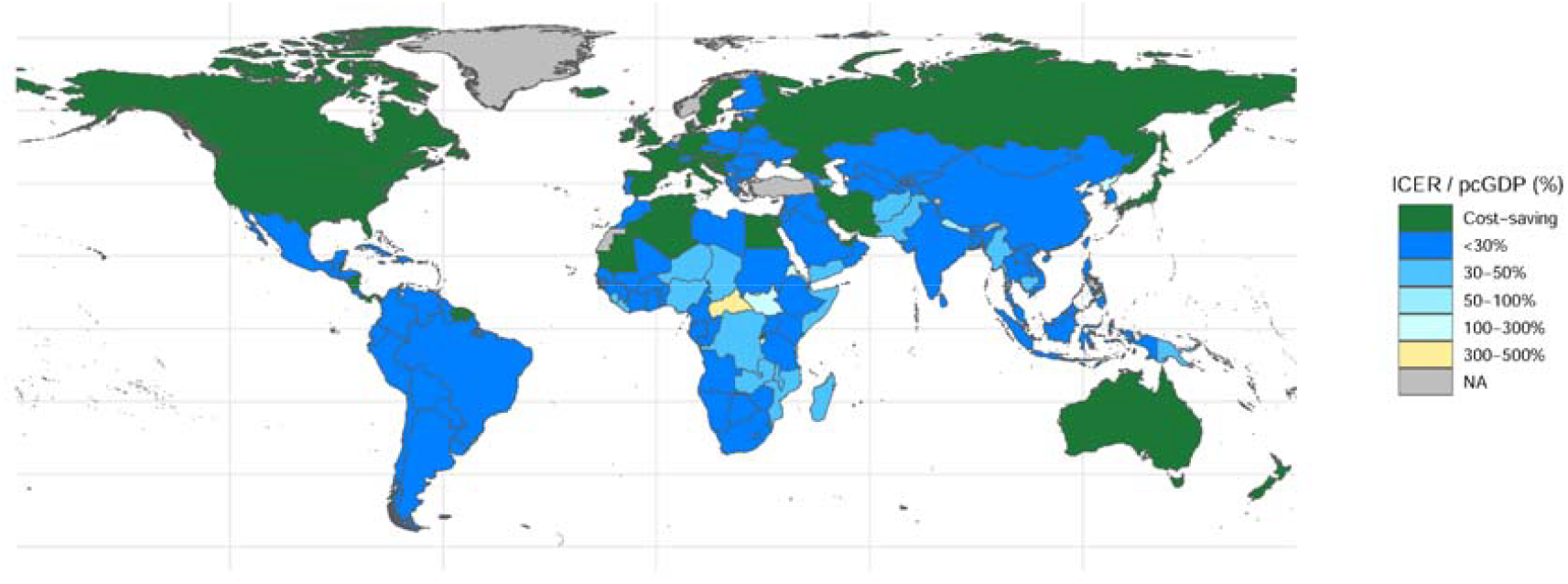
Incremental cost-effectiveness of adding preventive treatment without infection testing for contacts aged 15-44 years onto multidrug-resistant/rifampicin-resistant tuberculosis household contact management with preventive treatment for those under 15 years. Incremental Cost-Effectiveness Ratios (ICERs) are shown as a percentage of per capita national gross domestic product (pcGDP). Grey denotes missing data.

Global incremental outputs for 2024 compared to a baseline of no HHCM are shown in Figure 5, with cumulative numbers presented in Appendix section 3.0.2. Unlike ATT for rifampicin-susceptible tuberculosis, ATT for MDR/RR-tuberculosis increased as the increase in diagnoses from ACF outweighed the reduction in incidence from TPT. In aggregate, MDR-RR/HHCM with TPT for all would reduce tuberculosis incidence by 11,900 (10,400 to 13,800), and tuberculosis deaths by 9,960 (9,100 to 10,900). Restricting TPT to those with positive infection tests reduced averted tuberculosis incidence by 17.2% (11.5% to 22.2%) (see Appendix table A15). Adverse events increased with age, particularly after 45 years, with 165,000 (159,000 to 172,000) AEs and 71,000 (68,000 to 74,000) serious AEs (SAEs) predicted if offering TPT to all. 52.6% of SAEs in this total occur in those aged ≥45 years. Restricting TPT to those with positive infection tests reduced AEs by 20.8% (19.7% to 22.0%).

**Figure 5:**
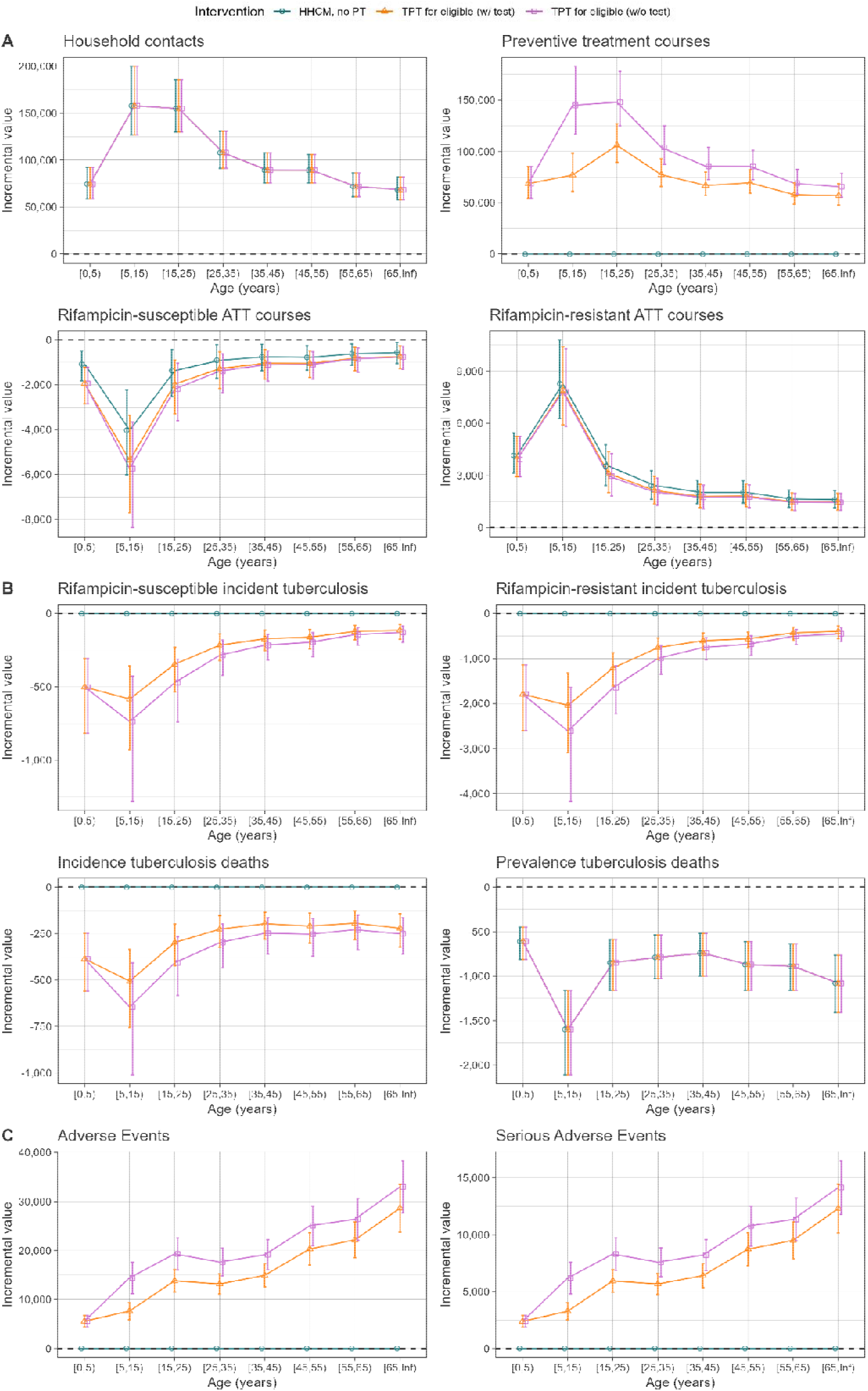
Global resource needs (A), health benefits (B), and adverse events (C) for multidrug-resistant/rifampicin-resistant tuberculosis household contact management by age group. Colours denote preventive treatment strategies. Dashed horizontal lines are at zero; scales vary by panel.

Global and regional incremental costs and DALYs compared to a baseline of no HHCM are shown in Appendix table A13. Use of TPT without testing across all ages would be associated with incremental costs of $209,000,000 ($161,000,000 to $275,000,000), and DALY reductions of 289,000 (262,000 to 321,000). With use of infection testing, incremental costs were 7.7% (4.4% to 10.8%) lower, and DALY reductions were 7.7% (4.9% to 10.2%) lower.

## Discussion

We found that extending MDR/RR-TPT use to older household contacts became less cost-effective with age. However, extending HHCM with TPT for contacts <15 years to include TPT for contacts aged up to 45 years was cost-effective in most countries, and cost-saving in 46/189 countries. In many countries, extending use of TPT to contacts aged over 45 years as part of MDR/RR-HHCM remained cost-effective, meaning that countries will need to consider additional factors such as affordability and capacity to manage AEs in making policy choices around age cut-offs. We found TPT without use of infection testing dominated strategies targeting TPT to contacts testing positive: moderate increases in costs and AEs were outweighed by larger increases in benefits. Globally, we found that increased global use of MDR/RR-TPT could avert substantial morbidity and mortality: MDR/RR-TPT in all ages without testing would have prevented up to 11,900 (10,500 to 13,800) tuberculosis episodes and 9,960 (9,080 to 10,900) deaths in 2024.

Our secondary analysis evaluating comparisons against a baseline of no MDR/RR-HHCM also demonstrated that the increase in diagnosis for prevalent MDR/RR-tuberculosis due to ACF outweighed the reduction in incidence via TPT. This is important for contexts where MDR/RR-HHCM is not occurring currently, as evaluation of contacts for prevalent disease following the identification of an individual with newly diagnosed MDR/RR-tuberculosis, without TPT, would avert substantial mortality and reduce transmission, even if provision of TPT is not possible.

Several cost-effectiveness analyses have considered household contact management for MDR/RR-tuberculosis and found interventions to be either cost-saving or generally likely to be cost-effective. A systematic review, published in 2017, evaluated five different MDR/RR-TPT regimens vs no TPT, finding four of them cost saving.^16^ Another study modelled fluoroquinolone TPT in the United States vs no TPT, finding that TPT was likely to lead to health system savings, and reduced mortality.^17^ Two global models, one before the results of the TB-CHAMP and VQUIN MDR trials were available, and one after, evaluated the impact of MDR/RR-HHCM and TPT on incident tuberculosis and mortality in children <15 years, finding these interventions cost-effective in most settings vs no MDR/RR-HHCM.^6,7^ An agent-based, household-structured tuberculosis and MDR/RR-tuberculosis transmission model was developed, calibrated to an India setting. Results from this model suggested substantial long-term population impact, but did not consider cost-effectiveness.^18^ These last three studies included impacts vs a comparator of no MDR/RR-HCM, and found the majority of benefits were from the case-finding component of MDR/RR-HCM. Finally, a cost-effectiveness analysis, conducted as part of the VQUIN MDR trial found that levofloxacin TPT cost-saving in Vietnam.^19^ Our results add to this body of work by extending the geographic range to a global context and evaluating all age groups.

In our analysis, the countries where extended use of TPT as part of MDR/RR-TPT proved cost-saving tended to be in the global north, where some of the highest proportions of MDR/RR-tuberculosis are found, and where resources are more likely to be available for expanded use of MDR/RR-TPT within existing programmes. However, the budgetary implications of wider use of MDR/RR-TPT are likely to be most important to countries with lower resources, where wider use of MDR/RR-TPT was less likely to be cost-saving. Cost-savings in richer countries are likely to be driven by the high baseline costs of managing tuberculosis disease, where preventing a single case yields substantial downstream savings in averted treatment costs.

We estimated that AE rates would escalate rapidly as older age groups were made eligible for TPT. Our definitions of AEs included arthritis or tendonitis, and many AEs will represent relatively minor inconveniences. However, SAEs could include tendon rupture, vascular aneurysms, or seizures,^20^ and should therefore be part of considerations in forming policy on TPT eligibility within MDR/RR-HHCM programmes. While we cost AEs due to treating incident tuberculosis, we did not separately estimate these numbers. However, the incremental number of AEs from ATT is likely to be smaller than from expanded TPT use because incidence occurs among only a fraction of contacts even without TPT. Programmes must plan for increased management of AEs if extending TPT use. More programme data on the nature of AEs and their dependence on age would be useful to refine policy as use of levofloxacin TPT scales up.

We considered a number of other outcomes dependent on age in our analysis. This included a novel analysis of the interpretation of a positive test for tuberculosis infection in contacts, and its relationship with community transmission intensity. For older ages and in higher transmission settings, positive tests of infection are less likely to represent recent (re)infection that carries higher risks of progression to tuberculosis disease. This aspect of the modelling may have wider application, and contributes to the lower cost-effectiveness of expanded TPT use in older age groups and higher transmission settings. This is despite a case-fatality ratio that was modelled as increasing with age.

Our analysis has primarily focused on age-specific TPT policies, however we were not able to parametrize age-dependence in all aspects of our analysis. This included the cost of hospitalisation, as well as the frequency of AEs for those on treatment. Wilkinson et al.^21^ show that age is an important determinant of hospitalization costs for paediatric MDR/RR-tuberculosis treatment in South Africa. We also did not fully consider transmission, which varies considerably by age. Although the indirect consequences of averted transmission chains in the future would be downweighted by discounting in cost-effectiveness analyses, including these effects would have increased the intervention impact and improved the cost-effectiveness. We also considered 100% coverage of interventions, which is unlikely to be achievable in a real-world setting, compared to a scenario with no intervention, which may not reflect current coverage.

In common with recent cost-effectiveness analyses, we include averted morbidity and mortality due to long-term effects in MDR/RR-tuberculosis survivors, which comprise a substantial burden of disease globally.^22^ However, we did not include post-tuberculosis costs to the health system, which have been treated as very substantial in some analyses.^19^ Furthermore, we did not consider patient costs. TPT can impose costs on households,^23^ but conversely has the potential to avert substantial treatment and post-tuberculosis-associated patient costs. Ongoing work on post-tuberculosis should clarify these economic consequences for patients.

Our results demonstrate extended use of TPT within MDR/RR-HHCM is often cost-effective, but that increasing age is associated with reduced cost-effectiveness and increasing AEs. While we do not demonstrate a clear age cut-off below which TPT should be provided and above which it should not, our results do suggest that universal TPT to all ages may not be the most appropriate approach.

Conversely, we find that TPT strategies using tuberculosis infection testing are dominated by those that do not, suggesting a limited role for testing in MDR/RR-HHCM. Even without TPT, the ACF component of MDR/RR-HHCM confers substantial health benefits on its own. Countries will need to weigh this evidence in deciding how to carry out MDR/RR-HHCM, which age groups to give TPT.

Choices should be informed by local tuberculosis epidemiology, economic constraints, current tuberculosis and other health programme considerations and workforce capacity.

## Data Availability

All code and data to reproduce this analysis are publicly available on GitHub https://github.com/nmafirakureva/DRHHCMextension

## Author contributions

All authors conceived and designed the study. PJD, NM and CFM did the data analysis and modelling. NM, PJD, CFM, MH and JS wrote the first draft of the Article. All authors contributed to writing the first draft of the Article. All authors designed the methods, critiqued the results, and revised and edited the submitted Article. NM and PJD directly accessed and verified the underlying data and had final responsibility for the decision to submit for publication.

