## Appendix for "Household contact management of drug-resistant tuberculosis: a global cost-effectiveness modelling analysis of expanded preventive treatment by age": HHDRextension_Appendix.docx

**1. Supplementary methods: epidemiological & outcomes model	2**

1.1. Index drug-resistance type & number of household contacts	2

1.1.1. Model for number of household contacts	2

1.1.2. Drug-resistance, HIV, and symptoms	5

1.2. Risks of infection, co-prevalent, and incident tuberculosis	6

1.2.1. Co-prevalence of tuberculosis disease	6

1.2.2. Tuberculosis infection and interpretation	6

1.2.3. Risks of progression to tuberculosis disease	7

1.3. Preventive treatment	8

1.3.1. Preventive treatment efficacy	8

1.3.2. Adverse events from preventive treatment	8

1.4. Tuberculosis treatment & outcomes	9

1.4.1. Case detection	9

1.4.2. Outcomes	9

1.5. Modelling approach	11

1.5.1. Parameter distributions	11

1.5.2. Computational approach & metrics calculated	12

1.5.3. Reproducibility	12

**2. Supplementary methods: costing	13**

2.1. Resource use	13

2.2. Unit costs	14

2.2.1. Sources & assumptions	14

2.2.2. Tuberculosis treatment costs	19

**3. Supplementary results	20**

3.0.1. Supplementary tables	20

3.0.2. Supplementary figures	38

**5. References	47**

### 1. Supplementary methods: epidemiological & outcomes model

#### 1.1. Index drug-resistance type & number of household contacts

##### 1.1.1. Model for number of household contacts

The number of household contacts in each of eight age groups (0-4, 5-14, 15-24, 25-34, 35-44, 45-55, 55-64 and over 65 years) living with adults (≥15 years) diagnosed with tuberculosis was estimated by extending a Bayesian multivariate regression model originally developed by Dodd and colleagues.^1^ The model used data obtained from the Demographic and Health Surveys (DHS) programme (<https://dhsprogram.com>) to predict age-stratified household composition based on: (1) individual characteristics (sex and age category: 15-24, 25-34, 35-44, 45-54, 55-64, or ≥65 years) and (2) country-level predictors from World Bank data (GDP per capita, life expectancy, infant mortality, population age structure, urbanization rate, population density, and total fertility rate). We maintained two key assumptions from prior analyses: (a) household structures of tuberculosis patients do not significantly differ from the general population, and (b) contact numbers are independent of drug-resistance status. The model was used to predict the number of household members of all ages in each country and age group for each of 189 countries.

DHS datasets were updated to the most recent survey years, and countries with newly available data were included (76 datasets compared to 69 in the previous analysis). (https://dhsprogram.com)

The countries included in this analysis were: Afghanistan, Albania, Angola, Armenia, Azerbaijan, Bangladesh, Benin, Bolivia, Brazil, Burkina Faso, Burundi, Cambodia, Cameroon, Cayman Islands, Central African Republic, Chad, Colombia, Comoros, Congo - Brazzaville, Congo - Kinshasa, Côte d’Ivoire, Dominican Republic, Egypt, Eswatini, Ethiopia, Gabon, Gambia, Ghana, Guam, Guinea, Guyana, Haiti, Honduras, India, Indonesia, Jordan, Kazakhstan, Kenya, Lebanon, Lesotho, Madagascar, Malawi, Maldives, Mali, Mauritania, Moldova, Morocco, Mozambique, Myanmar (Burma), Namibia, Nepal, New Caledonia, Nicaragua, Nigeria, Pakistan, Papua New Guinea, Paraguay, Peru, Philippines, Rwanda, Senegal, Sierra Leone, South Africa, São Tomé & Príncipe, Tajikistan, Tanzania, Timor-Leste, Togo, Turkey, Uganda, Ukraine, Uzbekistan, Vietnam, Yemen, Zambia, Zimbabwe

The DHS datasets were used to calculate the number of people in each age group living with individuals of specified age and sex within each household. The two figures below show the mean number of people in each age group living with individuals of given age and sex.

Mean number of people in each age group living with male individuals of given age.

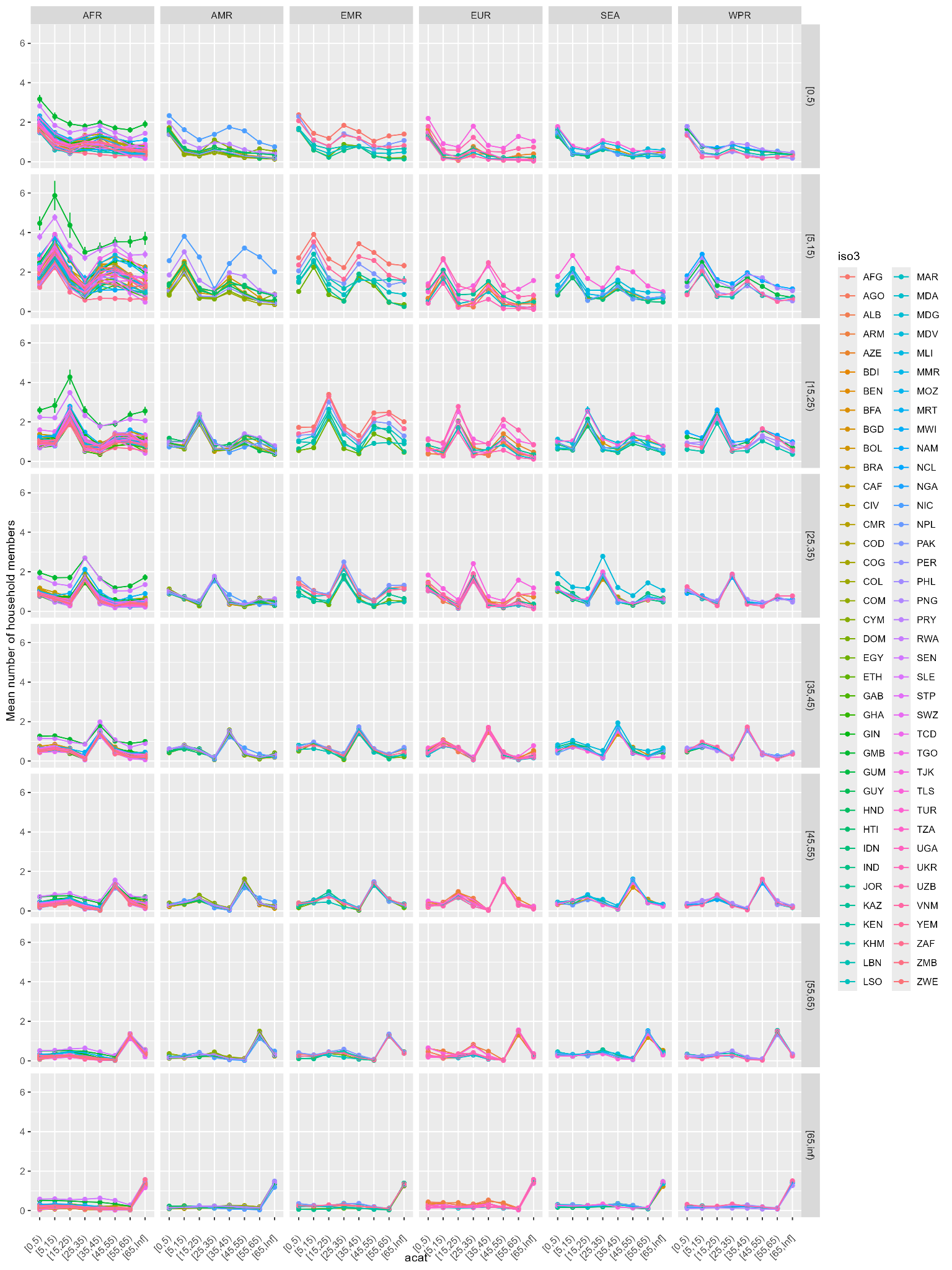

Mean number of people in each age group living with female individuals of given age.

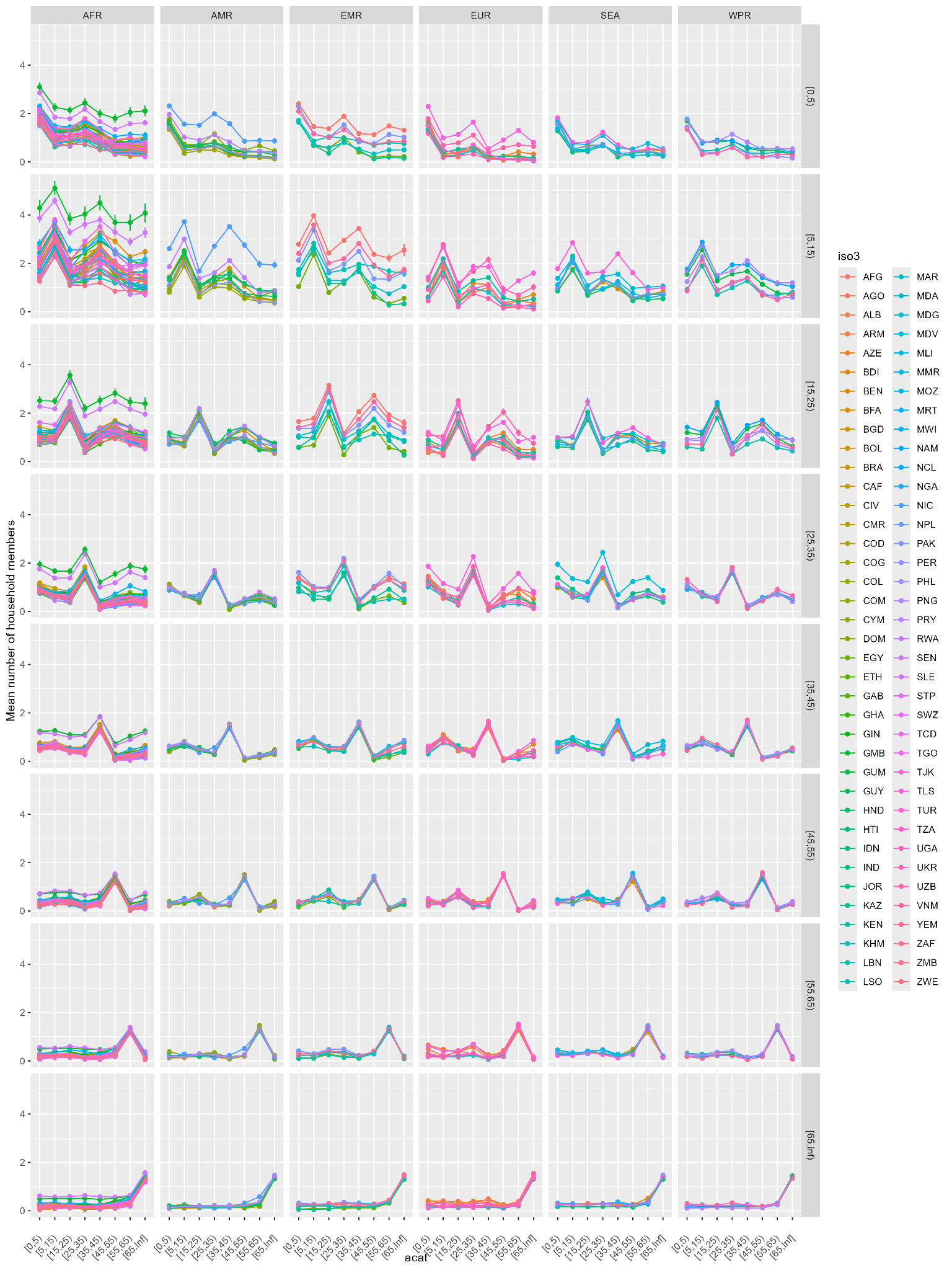

As in the previous analysis, we used World Health Organization (WHO) notification data with age- and sex-patterns for patients ≥15 years based on each country’s most recent data (2024). Because 2024 notifications and treatment data for RR/MDR-TB were missing, country-specific proportions relative to total TB notifications were calculated using complete 2019 baseline data and applied to 2024 total notifications to estimate missing values. Regional average patterns of the proportions were applied to total country notifications for countries with no age- and sex-stratified notification data. Similarly to a previous analysis,^2^ we assumed that all adult MDR/RR-TB notifications were pulmonary, since diagnosis of drug resistance commonly relies on bacteriological specimens, most often samples of pulmonary secretions. These age- and sex-specific notifications were combined with age- and sex-specific predictions of number of household members to estimate the average number of household contacts of each age category per tuberculosis patient in each country.

##### 1.1.2. Drug-resistance, HIV, and symptoms

We again used data from Chiang et al^3^ to model the concordance of drug resistance patterns, with a fraction 17.4% (9.1% to 27.7%) of household contacts being rifampicin-susceptible. For those contacts who concordantly have rifampicin resistance, we consider fluoroquinolone resistance prevalence based on country-level estimates of the proportion of MDR/RR-TB with fluoroquinolone resistance in Dodd et al.^2^

The WHO estimate of HIV prevalence among tuberculosis patients in each country was used for the proportion of index patients with HIV. As in Dodd et al.^2^, HIV prevalence in 0-4 year old and 5-14 year old household contacts of tuberculosis/HIV index was based on data in Martinez et al.^4^ For adults, HIV prevalence among all household contacts was found as 16.9% (14.7% to 19.1%) by Calderwood et al.^5^, and 25.2% (21.2% to 29.6%) among household TB/HIV contacts (their Appendix table L). They also observe an approximately 1.5x higher HIV prevalence in adults aged 40+ years compared to those aged 18 to 49 years (their Figure B). In order to model variation in HIV prevalence by country, we only model HIV in adult contacts of households where the index TB episode is living with HIV, but counterbalance this downward bias by applying the 1.5x factor to 35+ year age categories to the all-age 25.2% HIV prevalence estimate.

Antiretroviral therapy (ART) coverage for all groups was taken as the fraction of notified tuberculosis patients with HIV in the WHO data who were on ART.

The fraction of child contacts with symptoms is based on the discussion in Dodd et al.,^2^ which was based on Martinez et al^4^ and Sayedi et al.^6^ The fraction of adults with symptoms is taken to be the same as its value (19%) is very close to that observed in Calderwood et al.^5^ (20%).

#### 1.2. Risks of infection, co-prevalent, and incident tuberculosis

##### 1.2.1. Co-prevalence of tuberculosis disease

The co-prevalence of tuberculosis among child contacts of MDR/RR-TB is based on Shah et al.^7^ at 7.8% (5.6% to 10.0%). For adults, we use the meta-analysis of Velen et al.^8^, which found a co-prevalence for MDR/XDR index TB of 4.1% (2.8% to 5.6%),

##### 1.2.2. Tuberculosis infection and interpretation

As exposure to TB infection is cumulative, prevalence of evidence of TB infection due to background exposure increases with age. Correspondingly, the probability that a positive test for TB infection in a household contact implies recent infection, and therefore higher risks of progression to disease, decreases with age. We sought to capture this by modelling the probability by age that a positive test for TB infection is due to recent exposure.

We consider a 3-state compartmental model with constant flows between the states. *U* = uninfected*, R =* “recently” infected*,* and *L =* “distantly” infected. The notion of recent vs distant is defined by a timescale $1/r$ fixed at 2 years. We assume a background community hazard of infection $\lambda$, and a hazard ratio protection against reinfection conferred by previous infection, $v$. These assumptions correspond to the following ordinary differential equations (ODE):

$$U'= -\lambda U$$

$$R' =\lambda U - rR+v\lambda L$$

$$L'=rR-v\lambda L$$

where we assume that initially everyone is uninfected: $U(0)=1, R(0)=L(0) = 0$.

Adding the second two equations and writing $T=U+L$, gives

$$T'=\lambda U$$

We can solve the first ODE to give

$U=e^{-\lambda a}=1-T$.

We are interested in the proportion of those infected who were infected recently, $\rho=R/T$, and writing $\sigma=1-\rho$, this obeys the ODE

$\sigma'=L'/T-\sigma T'/T = r-\left[ r+v\lambda+\lambda U/T \right]\sigma=r-\left[ r+v\lambda+\lambda\frac{e^{-\lambda a}}{1-e^{-\lambda a}} \right]\sigma$.

We can solve this with an integrating factor to yield

$$\sigma(a,\lambda,v,r)=(1-e^{-\lambda a})^{-1}\left[ \frac{r}{\hat{r}}\left( 1-e^{-\hat{r}a} \right)-\frac{r}{\hat{r}-\lambda}\left( e^{-\lambda a}-e^{-\hat{r}a} \right) \right]$$

where $\hat{r}=r+v\lambda$.

For those who have recently shared a household with a person having TB disease, we assume an additional cumulative hazard of exposure $H = -log\left( 1-p_{H} \right)$.

The fraction of those with household exposure who were recently infected (including in the community) is $\hat{\rho}(a,\lambda,v,r,H)=A/B$ where the numerator is given by

$$A=e^{-\lambda a}\left( 1-e^{-H} \right)+\left( 1-e^{-\lambda a} \right)\left( 1-e^{-vH} \right)+\left( 1-e^{-\lambda a} \right)\sigma(a,\lambda,v,r)e^{-vH}$$

and the denominator by

$B=e^{-\lambda a}\left( 1-e^{-H} \right)+\left( 1-e^{-\lambda a} \right)$.

The terms in the numerator have the interpretation: (avoided community infection; received household infection) + (did not avoid community infection; received household reinfection) + (did not avoid community infection & was [re]infected recently; avoided household reinfection). The terms of the denominator have the interpretation: (avoided community infection; received household infection) + (did not avoid community infection).

The resulting proportion of household TB infection due to recent progression, and therefore at higher risk of progression, is shown in Figure 2 of the main manuscript.

##### 1.2.3. Risks of progression to tuberculosis disease

For children, we used the same approach to progression to disease described in Dodd et al.^2^ For adults, we used the age- and country-specific TB infection (TBI) estimates of Houben & Dodd^9^ to parametrize a background community force-of-infection, and combined it with the work in the previous section to parametrize a cumulative household infection hazard (based on child household contact TBI prevalence), a proportion of TBI among a given age group of household contacts that represents recent infection, and thus the probability of progression within 2 years. The probability of progression for recently-infected adults was based on Ragonnet et al.^10^

#### 1.3. Preventive treatment

##### 1.3.1. Preventive treatment efficacy

The effects of preventive treatment on progression to disease were parametrized as in Seddon et al.^11^

##### 1.3.2. Adverse events from preventive treatment

To capture the dependence of adverse even (AE) rates on age, we used a regression model of the dependence of AEs by age based on data in Yuen et al.^12^.

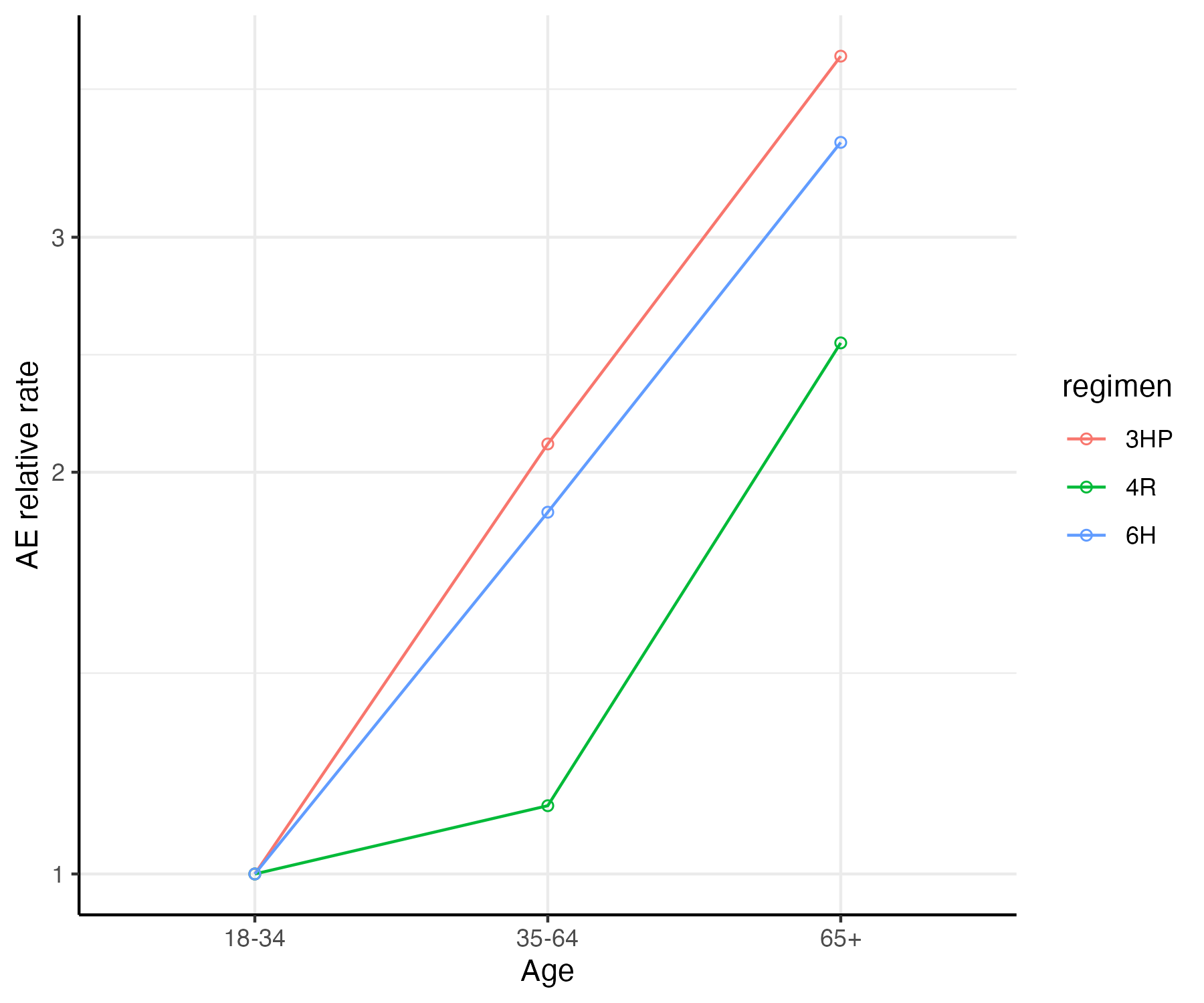

Figure A1 Adverse events by age from Yuen et al.^12^.

Fitting the log relative risk with linear regression have a dependence RR $\propto exp(0.0270\times age)$ (see Figure A1). Comparing the modelled distribution of AEs using this formula to the distribution seen in TB-CHAMP and VQUIN studies is shown in Table A1.

Table A1 Comparison of modelled vs observed AE rates by age

| **Age (years)** | **Data** | **Model** |
| --- | --- | --- |
| **0-2** | 0/224 (0%) | 6.5% |
| **3-4** | 3/184 (2%) | 6.9% |
| **5-9** | 0/29 (0%) | 7.6% |
| **10-17** | 10/83 (12%) | 9.1% |
| **18-29** | 43/204 (21%) | 11.9% |
| **30-44** | 60/268 (22%) | 17.1% |
| **45-59** | 81/313 (26%) | 25.7% |
| **60+** | 29/107 (27%) | 36.0% |

#### 1.4. Tuberculosis treatment & outcomes

##### 1.4.1. Case detection

Case detection was modelled in the same way as in Dodd et al, but extended to additional age groups.^2^ That is, we used WHO estimates of tuberculosis incidence in each country, sex, and age group together with notifications in the corresponding groups to calculate a community case detection ratio (CDR) for each group. As with previous work, we assumed a higher CDR for tuberculosis (co-prevalent or incident) among household contacts, to account for the household’s demonstrated access to care and experience with tuberculosis. In particular, we allowed the household CDR in each group to take an uncertain value between the background CDR for that group, and whichever is smaller of 1.5x the background and 1. To calculate the CDR for MDR/RR-TB, which we take as meaning the fraction of MDR/RR-TB receiving appropriate treatment, we used data on MDR/RR-TB treatment as the numerator and estimates of MDR/RR-TB incidence as the denominator. We assumed splits by age followed those seen in notification data for MDR/RR-TB notifications, and those seen in incidence estimates for MDR/RR-TB estimates.

##### 1.4.2. Outcomes

In order to capture the age-dependence of risks of death, we developed a unified representation of case-fatality ratios (CFRs) in children and adults. We based the pattern of CFR by age in both treated and untreated people with TB on data in two papers reporting on CFRs for people with TB on treatment: Osman et al.^13^ in children and adolescents, and Osman et al in adults.^14^ The 15-19 year age group was used as a reference category in the former, and taken to have the same risk as the 15-24 year age group in the latter. This was used to link data from the two papers for a relative risk of death across all ages (see Figure A2). The absolute CFRs in adults were scaled to produce an average on and off treatment that matches those used in WHO estimation processes, based on Tiemersma et al.^15^, and were represented by normal distributions on a logit scale (see table Table A2). Risks of death in paediatric age groups were modelled as an odds ratio applied to the treatment-specific risk of death in the 15-19 year age group; these odds ratios were represented by normal distributions on a logit scale(see table Table A3).

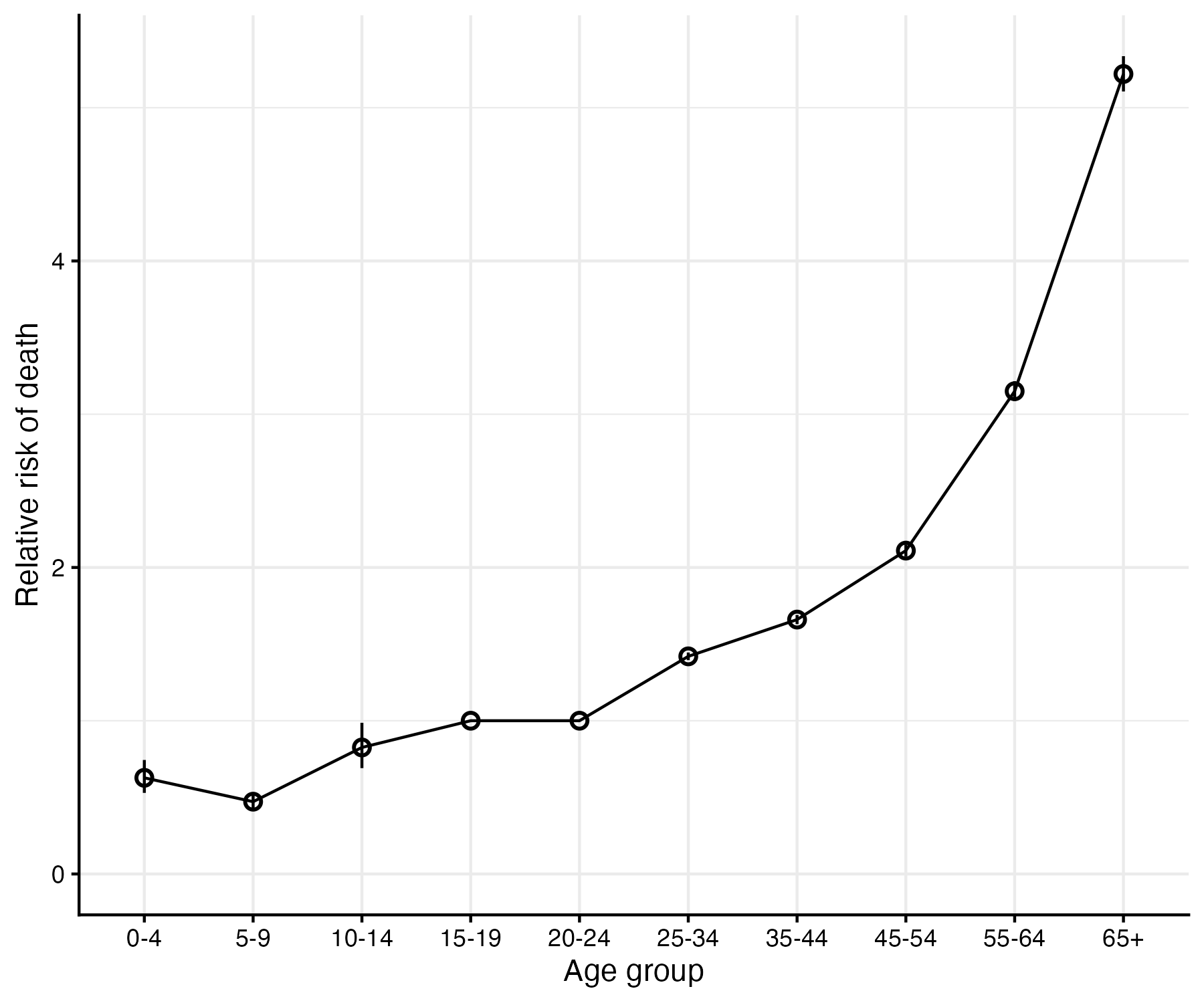

Figure A2 The relative risk of death for tuberculosis patients from two papers by Osman et al.

Table A2 The distributions used for logit (case fatality ratio) in adult age groups

| **ATT status** | **Age group** | **DISTRIBUTION** | **SOURCE** |
| --- | --- | --- | --- |
| **No treatment** | 15-24 | N(-0.89,0.02) | Osman 2021 |
| **No treatment** | 25-34 | N(-0.54,0.02) | Osman 2021 |
| **No treatment** | 35-44 | N(-0.39,0.02) | Osman 2021 |
| **No treatment** | 45-54 | N(-0.15,0.02) | Osman 2021 |
| **No treatment** | 55-64 | N(0.25,0.02) | Osman 2021 |
| **No treatment** | 65+ | N(0.76,0.02) | Osman 2021 |
| **On treatment** | 15-24 | N(-4.5,0.42) | Osman 2021 |
| **On treatment** | 25-34 | N(-4.15,0.42) | Osman 2021 |
| **On treatment** | 35-44 | N(-4.00,0.42) | Osman 2021 |
| **On treatment** | 45-54 | N(-3.76,0.42) | Osman 2021 |
| **On treatment** | 55-64 | N(-3.36,0.42) | Osman 2021 |
| **On treatment** | 65+ | N(-2.85,0.42) | Osman 2021 |

Table A3 The distributions used in paediatric age groups for logit(odds ratio) of death compared to the treatment-specific risk in the 15-19 year age group

| **Age group** | **DISTRIBUTION** | **SOURCE** |
| --- | --- | --- |
| **0-4** | N(-0.48,0.09) | Osman 2021b |
| **5-9** | N(-0.78,0.06) | Osman 2021b |
| **10-14** | N(-0.20,0.09) | Osman 2021b |

#### 1.5. Modelling approach

##### 1.5.1. Parameter distributions

The remaining parameters used to represent uncertainty in model parameters are shown in Table A4.

Table A4 Table of parameter distributions used in probabilistic sensitivity analysis and sources

| **NAME** | **DISTRIBUTION** | **DESCRIPTION** | **SOURCE** | **MEAN (IQR)** |
| --- | --- | --- | --- | --- |
| hivpi | LN(2.07,0.287) | IRR for TB incidence given HIV+/ART- (for individuals) | Dodd et al 2017 | 7.900 (6.540 - 9.543) |
| artp | LN(-1.27,0.157) | HR for TB incidence given HIV+/ART+ vs HIV+/ART- | Dodd et al 2016 | 0.300 (0.271 - 0.332) |
| HHhivprev04 | B(55,526) | Prevalence of HIV in child HH contacts of HIV+ index case | Martinez et al 2017 | 0.094 (0.086 - 0.103) |
| HHhivprev514 | B(54,854) | Prevalence of HIV in child HH contacts of HIV+ index case | Martinez et al 2017 | 0.059 (0.054 - 0.065) |
| HHhivprev15plus | B(103.19, 306.30) | Prevalence of HIV in adult HH contacts of HIV+ index case (x1.5 if 35+ years) | Calderwood et al 2024 | 0.252 (0.237 - 0.266) |
| LTBI04 | B(106.73,193.92) | LTBI prevalence | Fox et al 2013 | 0.355 (0.336 - 0.373) |
| LTBI514 | B(41.84,36.95) | LTBI prevalence | Fox et al 2013 | 0.531 (0.493 - 0.569) |
| LTBI04hi | B(10.62,54.55) | LTBI prevalence | Fox et al 2013 | 0.160 (0.131 - 0.192) |
| LTBI514hi | B(17.04,75.56) | LTBI prevalence | Fox et al 2013 | 0.182 (0.156 - 0.210) |
| iptRR | B(1.82,2.98) | RR for incident TB given Lfx, age <15 | TB-CHAMP | 0.380 (0.220 – 0.523) |
| iptRRtstpos | B(3.14,31.53) | RR for incident TB given PT in TST+, age <15 | Martinez et al 2018 ratio applied to TB-CHAMP | 0.091 (0.055 - 0.118) |
| iptRRhivpos | B(1.82,2.98) | RR for incident TB given PT in HIV+, age <15 | TB-CHAMP | 0.380 (0.220 – 0.523) |
| CFRrtx.RR | B(11.09,127.53) | CFR RR-ATT when truly RR | Harausz et al 2018 | 0.078 (0.064 - 0.094) |
| coprevDRkids | LN(2.05,0.14) | coprevalence in percent of HH contacts of DRTB, children | Shah et al 2017 | 7.800 (7.089 - 8.583) |
| coprevDRadults | LN(1.41,0.17) | coprevalence in percent of HH contacts of DRTB, adults | Velen et al 2021 | 4.100 (3.655 - 4.600) |
| concord | B(57.40,12.09) | concordance in DR type | Chiang 2020 | 0.829 (0.797 - 0.858) |
| prog04 | B(5.15,21.97) | LTBI+ progression u5 | Martinez et al 2018 | 0.182 (0.136 - 0.236) |
| prog514 | B(4.15,43.02) | LTBI+ progression u5 | Martinez et al 2018 | 0.082 (0.058 - 0.112) |
| fracSymptomatic | B(2.94,11.74) | Fraction of contacts presumptive TB | Martinez 2017 & Sayedi 2020 | 0.186 (0.124 - 0.262) |
| fracAE | B(226,1186) | Fraction of PT any MS AE | see Appendix | 0.160 (0.153 - 0.167) |
| fracSAE | B(97,1315) | Fraction of PT G2+ MS AE | see Appendix | 0.068 (0.064 - 0.073) |
| prog15plus | LN(-2.84,0.32) | Progression risk from recent infection (15+ years) | Ragonnet et al 2017 | 0.062 (0.031 – 0.110) |

##### 1.5.2. Computational approach & metrics calculated

We used the same decision tree structure for clinical pathways and outcomes as in Dodd et al.,^2^ with modifications to the data representing the cohorts entering the tree and the parametrization of costs and probabilities. We again used 1,000 samples for all parameter distributions, including country-level fluroquinolone resistance.

We calculated measures of resource use, such as anti-tuberculosis treatment (stratified by regimen type) and preventive therapy outcomes, and tuberculosis disease outcomes. Life-years lost were calculated based on age groups and they discounted contribution to DALYs associated with each strategy calculated. The contribution to DALYs from morbidity during and after tuberculosis were included by using a decrement to the quality of life in TB survivors of 0.036 based on the median in Menzies et al.,^16^ and a causal mortality hazard ratio of 1.78 in survivors from Menzies et al.^16^

##### 1.5.3. Reproducibility

Code and data to run this analysis can be found on GitHub at <https://github.com/nmafirakureva/DRHHCMextension>

### 2. Supplementary methods: costing

#### 2.1. Resource use

We followed the same approach described in Dodd et al.^2^, to estimate resource use and costs from a health system perspective. The cost analysis was extended to include resource use and costs for adults aged 15 years and older. In brief, the analysis focussed on costs of resources used for household visits for screening, TB infection testing, tuberculosis testing, treatment of tuberculosis disease, and provision of TPT. Table A5 provides a summary of HHCM activities and associated resource utilisation for the different intervention scenarios. We assumed a single home visit for tuberculosis symptomatic screening, an HIV test for non-symptomatic contacts aged ≥5 years, a tuberculin skin test (TST) for non-symptomatic contacts aged ≥5 years and 6 months of daily TPT provision. TPT provision comprised resources such as levofloxacin drugs, monthly health facility follow-up visits, monthly laboratory monitoring (complete blood count and liver function tests) and management of adverse events experienced during TPT. Similar assumptions on dosing for children and management of adverse effects were followed.

Table A5 Activities involved in each costing each intervention. HIV = human immunodeficiency virus; Lfx = Levofloxacin; TPT = tuberculosis preventive treatment; TST = Tuberculin Skin Test.

| **Intervention** | | | | |
| --- | --- | --- | --- | --- |
| Household contact management | No | Yes | | |
| TPT recipients | None | None | <5 & HIV+ & TST+ | All contacts |
| TPT regimen | None | None | Lfx | Lfx |
| **Household contact management activities** | | | | |
| Home visit |  | ✓ | ✓ | ✓ |
| HIV test for ≥ 5 years |  | ✓ | ✓ |  |
| TST for ≥ 5 years |  |  |  |  |
| **Tuberculosis testing if contact found symptomatic** | | | | |
| Outpatient department visit |  | ✓ | ✓ | ✓ |
| Chest X-ray |  | ✓ | ✓ | ✓ |
| Xpert MTB/RIF |  | ✓ | ✓ | ✓ |
| **TPT** | | | | |
| Drug costs |  |  | ✓ | ✓ |
| Follow up |  |  | ✓ | ✓ |
| Monitoring |  |  | ✓ | ✓ |
| Adverse events |  |  | ✓ | ✓ |
| **Anti-TB treatment** | ✓ | ✓ | ✓ | ✓ |

Similarly, we assumed symptomatic contacts would require a single outpatient facility visit, a chest X-ray and an Xpert MTB/RIF test on a single sputum sample. Younger children aged 0-4 years would require an induced sputum sample. All contacts diagnosed with tuberculosis would be initiated in anti-tuberculosis treatment and accumulate associated costs as described in Dodd et al for children. For adults, resource use for anti-tuberculosis treatment was based on 2024 country reported data in the WHO GTB database. Table A6 summarises the resources used for the treatment of tuberculosis disease.

Table A6 Summary of tuberculosis testing and treatment resource utilisation, independent of intervention. RS-TB=drug-susceptible tuberculosis; HIV = human immunodeficiency virus; MDR/RR-TB=multidrug- or rifampicin-resistant tuberculosis; RS-TB=drug-susceptible tuberculosis; B=bedaquiline; D=Delaminid; L=Linezolind; C=Clofazimine; Lfx=Levofloxacin; H=isoniazid; R=rifampicin; Z=Pyrazinamide; E=ethambutol; NTP= National Tuberculosis Program

| **Activity** | **Resources** | **Resource description** |
| --- | --- | --- |
| MDR/RR-TB treatment | HIV test | Health facility-based HIV testing per testing episode/person. |
|  | Inpatient care | Inpatient care (hospitalisation) for the entire MDR/RR-TB episode per patient. |
|  | Outpatient care | Outpatient care (health facility treatment follow-up visits) for the entire MDR/RR-TB episode per patient. |
|  | NTP programme costs | NTP programme costs per patient starting MDR-TB/XDR-TB treatment. |
|  | Drugs | Children 0-14 years with fluoroquinolone resistant MDR/RR-TB: 9 months BDLC. |
|  |  | Children 0-14 years with fluoroquinolone susceptible MDR/RR-TB: 9 months BDLLfx. |
|  |  | Adults 15 years and over with fluoroquinolone resistant MDR/RR-TB: 6 months BPaL. |
|  |  | Adults 15 years and over with fluoroquinolone susceptible MDR/RR-TB: 6 months BPaLM. |
|  | Laboratory monitoring | Children < 15 years: full blood count and electrocardiogram every 2 months for 9 months. |
|  |  | Adults 15 years and over: full blood count and electrocardiogram every 2 months for 6 months. |
| RS-TB treatment | HIV test | Health facility-based HIV testing per testing episode/person. |
|  | Inpatient care | Inpatient care (hospitalisation) for the entire RS-TB episode per patient. |
|  | Outpatient care | Outpatient care (health facility treatment follow-up visits) for the entire RS-TB episode per patient. |
|  | NTP programme costs | NTP programme costs per patient starting first-line tuberculosis treatment. |
|  | Drugs | 2 months H-R-Z-E + 4 months H-R for children. |
|  |  | Typical country-level first-line drugs for adults. |
|  | Laboratory monitoring | Assumed no laboratory monitoring during treatment. |

#### 2.2. Unit costs

##### 2.2.1. Sources & assumptions

All unit costs used in this analysis were estimated using publicly available data or published literature (see Table A7) following the approach in Dodd et al. For adults, costs were based on country reported costs in the WHO GTB database with regression analysis used to predict costs for countries with missing data. Table Table A6 summarises the resources used for the treatment of tuberculosis disease. Unit costs were converted to 2024 USD prices using standard adjustments. Uncertainty was accounted for by modelling all unit costs as following gamma distributions with means/SDs corresponding to estimates.

Table A7 Unit costs of tuberculosis screening, tuberculosis testing and diagnosis, tuberculosis treatment and LTBI treatment.

| **Resource** | **Cost parameter** | **Description** | **Unit cost ($US)** | **Unit cost parameter estimation assumptions** |
| --- | --- | --- | --- | --- |
| Household visit | c_hh_visit | Household visit to perform active TB contact tracing and screening. | 3.36 (0.8) | [Estimated based on costs for active case finding per household contact reported in a study](https://doi.org/10.1016/j.ijid.2020.09.034) from Peru. |
| HIV test | c_hiv_test | Facility-based HIV testing | Income-group specific | [Estimated based on income level group (low- to upper-middle income countries versus high income countries) specific health facility-based HIV testing costs per testing episode from](https://www.ncbi.nlm.nih.gov/books/NBK316032/). |
| Tuberculin skin test | c_tst_test | Tuberculin skin test | 7.26 (1.9) | [Estimated based on costs for a tuberculin skin test with purified protein derivative (PPD RT 23) reported in a recent study](https://www.ncbi.nlm.nih.gov/pmc/articles/PMC7733491/) from Brazil. |
| QuantiFERON-TB Gold Plus (QFT-Plus) test | c_tst_test | QuantiFERON-TB Gold Plus (QFT-Plus) test | 21.02 (5.3) | [Estimated based on costs for QuantiFERON-TB Gold Plus (QFT-Plus) test reported in a recent study](https://www.ncbi.nlm.nih.gov/pmc/articles/PMC7733491/) from Brazil. |
| Outpatient visit | c_opd_visit | Outpatient visit to a primary-level hospital in the public sector | Country-specific | [Estimated using a WHO CHOosing Interventions that are Cost-Effective (WHO-CHOICE) based econometric model](https://resource-allocation.biomedcentral.com/articles/10.1186/s12962-018-0095-x) using the number of total outpatient visits per facility per year, visits per provider per day and GDP per capita as predictors. |
| Chest X-ray | c_cxr_exam | Chest radiograph or chest X-ray | 4.04 (1.1) | [Estimated based on costs for a chest radiograph reported in a recent study](https://www.ncbi.nlm.nih.gov/pmc/articles/PMC7733491/) from Brazil. |
| Xpert MTB/RIF test (0-4 years) | c_xpert_test.04 | GeneXpert MTB/RIF molecular test in children below 5 years | 24.1 (2.1) | Comprises the costs for collecting an induced sputum sample (assuming inability to spontaneously produce sputum in young child) based on estimates provided by the Paediatric Operational Sustainability Expertise Exchange group ([POSEE](https://www.dropbox.com/sh/rmzlqoot9m1muxe/AACOPh8SuS4WwMQD_vfN-oHCa?dl=0) group) and the costs for a GeneXpert test on a single sample reported in a multi-laboratory [study](http://www.ncbi.nlm.nih.gov/pmc/articles/pmc6443160/) from India. |
| Xpert MTB/RIF test (>=15 years) | c_xpert_test.O15 | GeneXpert MTB/RIF molecular test in peo over 5 years | 17.5 (0.8) | Comprises the costs for collecting self-expectorated sputum based on recent [estimates](http://www.ncbi.nlm.nih.gov/pmc/articles/pmc7927348/) from Uganda and the costs for a GeneXpert test on a single sample reported in a multi-laboratory [study](http://www.ncbi.nlm.nih.gov/pmc/articles/pmc6443160/) from India. |
| Isoniazid-based TPT (0-4 years) | c_tpt_INH.04 | Isoniazid-based TB preventive treatment in children below 5 years | 5.4 (1.4) | [Estimated using weight band based dosing using the median weight for children in the 0-4 years age group and applying unit costs available from the Global Drug Facility](http://www.stoptb.org/gdf/) for a 6 month treatment on Isoniazid-based TPT. |
| Isoniazid-based TPT (5-14 years) | c_tpt_INH.514 | Isoniazid-based TB preventive treatment in children over 5 years | 12.6 (2.3) | [Estimated using weight band based dosing using the median weight for children in the 5-14 years age group and applying unit costs available from the Global Drug Facility](http://www.stoptb.org/gdf/) for a 6 month treatment on Isoniazid-based TPT. |
| Levofloxacin-based TPT (0-4 years) | c_tpt_LVX.04 | Levofloxacin-based TB preventive treatment in children below 5 years | 34.2 (11.7) | [Estimated using weight band based dosing using the median weight for children in the 0-4 years age group and applying unit costs available from the Global Drug Facility](http://www.stoptb.org/gdf/) for a 6 month treatment on Levofloxacin-based TPT. |
| Levofloxacin-based TPT (5-14 years) | c_tpt_LVX.514 | Levofloxacin-based TB preventive treatment in children over 5 years | 46.8 (22.9) | [Estimated using weight band based dosing using the median weight for children in the 5-14 years age group and applying unit costs available from the Global Drug Facility](http://www.stoptb.org/gdf/) for a 6 month treatment on Levofloxacin-based TPT. |
| Levofloxacin-based TPT (>=15 years) | c_tpt_LVX.O15 | Levofloxacin-based TB preventive treatment in adults (>=15 years) | Country-specific | Estimated using the average cost of drugs budgeted per patient for TB preventive treatment available from the WHO Global TB data |
| TPT follow-up | c_tpt_fu | TB preventive treatment follow-up | Country-specific |  |
| Isoniazid-based TPT monitoring | c_monit_INH | Isoniazid-based TB preventive treatment monitoring | Country-specific | Comprises the costs for collecting an induced sputum sample (assuming inability to spontaneously produce sputum in young child) based on estimates provided by the Paediatric Operational Sustainability Expertise Exchange group ([POSEE](https://www.dropbox.com/sh/rmzlqoot9m1muxe/AACOPh8SuS4WwMQD_vfN-oHCa?dl=0) group) and the costs for a GeneXpert test on a single sample reported in a multi-laboratory [study](http://www.ncbi.nlm.nih.gov/pmc/articles/pmc6443160/) from India. |
| Fluoroquinolone-based TPT monitoring | c_monit_FQ | Fluoroquinolone-based TB preventive treatment monitoring | Country-specific |  |
| Isoniazid-based TPT mild adverse events treatment | c_aes_INH | Isoniazid-based TPT mild adverse events treatment | Country-specific | The cost of treating children experiencing mild adverse events while receiving TPT and comprises one‐time outpatient visit and laboratory testing (complete blood count, electrolyte panel, urinalysis and liver function tests). The outpatient visit cost was estimated using a [WHO-CHOICE based econometric model](http://www.ncbi.nlm.nih.gov/pmc/articles/pmc156022/) as described before. Laboratory testing costs were estimated based on costs for LFTs and CBC reported in a [study from Brazil](https://www.ncbi.nlm.nih.gov/pmc/articles/PMC7733491/). |
| Fluoroquinolone-based TPT mild adverse events treatment | c_aes_FQ | Fluoroquinolone-based TPT mild adverse events treatment | Country-specific |  |
| Isoniazid-based TPT serious adverse events treatment | c_saes_INH | Isoniazid-based TPT serious adverse events treatment | Country-specific | The cost of treating children experiencing serious adverse events while receiving TPT and comprises 7 day hospitalisation and laboratory testing (complete blood count, electrolyte panel, urinalysis and liver function tests). The inpatient cost was estimated using a [WHO-CHOICE based econometric model](http://www.ncbi.nlm.nih.gov/pmc/articles/pmc156022/) as described before. Laboratory testing costs were estimated based on costs for LFTs and CBC reported in a [study from Brazil](https://www.ncbi.nlm.nih.gov/pmc/articles/PMC7733491/). |
| Fluoroquinolone-based TPT serious adverse events treatment | c_saes_FQ | Fluoroquinolone-based TPT serious adverse events treatment | Country-specific |  |
| DS-TB treatment (0-14 years) | c_dstb_tx.014 | Treatment for drug-susceptible TB in children below 15 years | Country-specific | Estimated using publicly available data based on an approach described in the WHO Global TB report as the sum of NTP expenditures, total costs for use of inpatient and outpatient care and cost of [paediatric specific anti-TB drugs](http://www.stoptb.org/gdf/). Two drug regimens were considered, based on fluoroquinolone resistance. |
| MDR/RR-TB treatment (0-14 years) | c_mdrtb_tx.014 | Treatment for multi-drug resistant/rifampicin resistant TB in children 0-14 years | Country-specific |  |
| DS-TB treatment (>=15 years) | c_dstb_tx.O15 | Treatment for drug-susceptible TB in adults (over 15 years) | Country-specific | Estimated using publicly available data based on an approach described in the WHO Global TB report as the sum of NTP expenditures, total costs for use of inpatient and outpatient care and cost of first-line and second-line drugs.Two drug regimens were considered, based on fluoroquinolone resistance. |
| MDR/RR-TB treatment (>=15 years) | c_mdrtb_tx.O15 | Treatment for multi-drug resistant/rifampicin resistant TB in adults (over 15 years) | Country-specific |  |

##### 2.2.2. Tuberculosis treatment costs

Tuberculosis disease treatment costs were estimated using the most recent publicly available data based on an approach described in the WHO Global Tuberculosis Report 2019^17^ and previously applied in Dodd et al.^2^ Total costs are calculated as the sum of NTP expenditures, costs for inpatient and outpatient care and the cost of anti-tuberculosis drugs (first-line or second-line). For children under 15 years, we used the cost of paediatric specific anti-tuberculosis drugs estimated based on unit costs available from the Global Drug Facility.^18^ Following the most recent treatment guidance, children (0–14 years) with MDR/RR-TB received 9 months of BDLC if fluoroquinolone-resistant or 9 months of BDLLfx if fluoroquinolone-susceptible.^19,20^ Adults (15 years and over) received 6 months of BPaL if fluoroquinolone-resistant or 6 months of BPaLM if fluoroquinolone-susceptible.^19^

### 3. Supplementary results

##### 3.0.1. Supplementary tables

Table A8 Resource use associated with full coverage of household contact management in each contact age group. Tuberculosis preventive treatment (PT) is assumed to be offered to those eligible. RS = Rifampicin-susceptible; RR = Rifampicin-resistant.

| **Age** | **Intervention** | **Household contacts** | **PT courses** | **RR-ATT** | **RS-ATT** |
| --- | --- | --- | --- | --- | --- |
| [0,5) | HHCM, no PT_None | 74,500 (58,600 - 92,500) | 0 (0 - 0) | 4,160 (3,130 - 5,340) | -1,080 (-1,790 - -458) |
| [0,5) | TPT for eligible (w/ test)_Lfx | 74,500 (58,600 - 92,500) | 68,600 (54,000 - 85,300) | 3,920 (2,900 - 5,110) | -1,930 (-2,750 - -1,200) |
| [0,5) | TPT for eligible (w/o test)_Lfx | 74,500 (58,600 - 92,500) | 68,600 (54,000 - 85,300) | 3,920 (2,900 - 5,110) | -1,930 (-2,750 - -1,200) |
| [5,15) | HHCM, no PT_None | 164,000 (138,000 - 199,000) | 0 (0 - 0) | 8,340 (6,910 - 10,200) | -5,380 (-7,260 - -3,830) |
| [5,15) | TPT for eligible (w/ test)_Lfx | 164,000 (138,000 - 199,000) | 80,100 (67,100 - 98,900) | 7,930 (6,530 - 9,810) | -7,030 (-9,230 - -5,370) |
| [5,15) | TPT for eligible (w/o test)_Lfx | 164,000 (138,000 - 199,000) | 151,000 (127,000 - 184,000) | 7,820 (6,380 - 9,750) | -7,460 (-9,790 - -5,720) |
| [15,25) | HHCM, no PT_None | 160,000 (143,000 - 181,000) | 0 (0 - 0) | 3,160 (2,630 - 3,800) | -2,040 (-2,610 - -1,480) |
| [15,25) | TPT for eligible (w/ test)_Lfx | 160,000 (143,000 - 181,000) | 110,000 (98,400 - 124,000) | 2,670 (2,100 - 3,330) | -2,770 (-3,440 - -2,150) |
| [15,25) | TPT for eligible (w/o test)_Lfx | 160,000 (143,000 - 181,000) | 153,000 (137,000 - 173,000) | 2,490 (1,880 - 3,160) | -3,040 (-3,770 - -2,390) |
| [25,35) | HHCM, no PT_None | 111,000 (102,000 - 122,000) | 0 (0 - 0) | 2,690 (2,030 - 3,530) | -831 (-1,480 - -153) |
| [25,35) | TPT for eligible (w/ test)_Lfx | 111,000 (102,000 - 122,000) | 80,200 (73,100 - 87,900) | 2,470 (1,790 - 3,340) | -1,180 (-1,830 - -503) |
| [25,35) | TPT for eligible (w/o test)_Lfx | 111,000 (102,000 - 122,000) | 107,000 (97,500 - 117,000) | 2,400 (1,720 - 3,280) | -1,290 (-1,960 - -598) |
| [35,45) | HHCM, no PT_None | 92,500 (85,300 - 101,000) | 0 (0 - 0) | 2,110 (1,530 - 2,780) | -586 (-1,080 - -116) |
| [35,45) | TPT for eligible (w/ test)_Lfx | 92,500 (85,300 - 101,000) | 69,800 (64,200 - 76,300) | 1,900 (1,280 - 2,540) | -830 (-1,350 - -330) |
| [35,45) | TPT for eligible (w/o test)_Lfx | 92,500 (85,300 - 101,000) | 88,600 (81,700 - 96,700) | 1,850 (1,230 - 2,490) | -891 (-1,420 - -382) |
| [45,55) | HHCM, no PT_None | 92,200 (83,900 - 102,000) | 0 (0 - 0) | 2,050 (1,470 - 2,730) | -561 (-1,060 - -51.8) |
| [45,55) | TPT for eligible (w/ test)_Lfx | 92,200 (83,900 - 102,000) | 72,300 (65,700 - 79,500) | 1,850 (1,270 - 2,540) | -783 (-1,280 - -264) |
| [45,55) | TPT for eligible (w/o test)_Lfx | 92,200 (83,900 - 102,000) | 88,400 (80,300 - 97,300) | 1,810 (1,210 - 2,510) | -827 (-1,340 - -304) |
| [55,65) | HHCM, no PT_None | 74,100 (67,800 - 82,000) | 0 (0 - 0) | 1,620 (1,150 - 2,190) | -415 (-791 - -41.8) |
| [55,65) | TPT for eligible (w/ test)_Lfx | 74,100 (67,800 - 82,000) | 60,000 (54,700 - 66,500) | 1,460 (1,000 - 2,030) | -578 (-969 - -185) |
| [55,65) | TPT for eligible (w/o test)_Lfx | 74,100 (67,800 - 82,000) | 71,000 (65,000 - 78,400) | 1,430 (960 - 2,000) | -604 (-1,000 - -203) |
| [65,Inf) | HHCM, no PT_None | 70,400 (64,300 - 77,000) | 0 (0 - 0) | 1,480 (993 - 2,030) | -335 (-635 - -5.38) |
| [65,Inf) | TPT for eligible (w/ test)_Lfx | 70,400 (64,300 - 77,000) | 59,100 (53,300 - 65,800) | 1,320 (835 - 1,890) | -477 (-795 - -154) |
| [65,Inf) | TPT for eligible (w/o test)_Lfx | 70,400 (64,300 - 77,000) | 67,400 (61,600 - 74,000) | 1,300 (809 - 1,860) | -494 (-820 - -171) |

Table A9 Incremental outcomes associated with full coverage of household contact management in each contact age group. Tuberculosis preventive treatment is assumed to be offered to those eligible. Lfx = Levofloxacin; RS = Rifampicin-susceptible; RR = Rifampicin-resistant.

| **Age** | **Intervention** | **Incident TB episodes** | **Incident RS-TB episodes** | **Incident RR-TB episodes** | **Incident TB deaths** | **Prevalent TB deaths** |
| --- | --- | --- | --- | --- | --- | --- |
| [0,5) | HHCM, no PT_None | 0 (0 - 0) | 0 (0 - 0) | 0 (0 - 0) | 0 (0 - 0) | -611 (-816 - -449) |
| [0,5) | TPT for eligible (w/ test)_Lfx | -2,270 (-3,190 - -1,540) | -509 (-827 - -296) | -1,770 (-2,550 - -1,110) | -382 (-546 - -240) | -611 (-816 - -449) |
| [0,5) | TPT for eligible (w/o test)_Lfx | -2,270 (-3,190 - -1,540) | -509 (-827 - -296) | -1,770 (-2,550 - -1,110) | -382 (-546 - -240) | -611 (-816 - -449) |
| [5,15) | HHCM, no PT_None | 0 (0 - 0) | 0 (0 - 0) | 0 (0 - 0) | 0 (0 - 0) | -1,550 (-1,950 - -1,240) |
| [5,15) | TPT for eligible (w/ test)_Lfx | -2,700 (-3,780 - -1,970) | -597 (-1,020 - -383) | -2,100 (-3,040 - -1,440) | -497 (-686 - -353) | -1,550 (-1,950 - -1,240) |
| [5,15) | TPT for eligible (w/o test)_Lfx | -3,410 (-4,950 - -2,370) | -758 (-1,310 - -456) | -2,650 (-4,070 - -1,730) | -627 (-920 - -423) | -1,550 (-1,950 - -1,240) |
| [15,25) | HHCM, no PT_None | 0 (0 - 0) | 0 (0 - 0) | 0 (0 - 0) | 0 (0 - 0) | -720 (-866 - -592) |
| [15,25) | TPT for eligible (w/ test)_Lfx | -1,510 (-1,970 - -1,170) | -334 (-502 - -223) | -1,180 (-1,600 - -868) | -260 (-350 - -195) | -720 (-866 - -592) |
| [15,25) | TPT for eligible (w/o test)_Lfx | -2,070 (-2,680 - -1,600) | -457 (-691 - -302) | -1,610 (-2,210 - -1,180) | -356 (-479 - -265) | -720 (-866 - -592) |
| [25,35) | HHCM, no PT_None | 0 (0 - 0) | 0 (0 - 0) | 0 (0 - 0) | 0 (0 - 0) | -888 (-1,180 - -672) |
| [25,35) | TPT for eligible (w/ test)_Lfx | -948 (-1,220 - -723) | -210 (-325 - -142) | -737 (-987 - -537) | -242 (-345 - -171) | -888 (-1,180 - -672) |
| [25,35) | TPT for eligible (w/o test)_Lfx | -1,250 (-1,600 - -951) | -277 (-426 - -187) | -971 (-1,310 - -706) | -319 (-454 - -223) | -888 (-1,180 - -672) |
| [35,45) | HHCM, no PT_None | 0 (0 - 0) | 0 (0 - 0) | 0 (0 - 0) | 0 (0 - 0) | -784 (-1,030 - -568) |
| [35,45) | TPT for eligible (w/ test)_Lfx | -741 (-942 - -588) | -164 (-243 - -111) | -576 (-780 - -437) | -193 (-281 - -135) | -784 (-1,030 - -568) |
| [35,45) | TPT for eligible (w/o test)_Lfx | -929 (-1,190 - -737) | -206 (-305 - -138) | -723 (-983 - -547) | -242 (-351 - -167) | -784 (-1,030 - -568) |
| [45,55) | HHCM, no PT_None | 0 (0 - 0) | 0 (0 - 0) | 0 (0 - 0) | 0 (0 - 0) | -900 (-1,190 - -641) |
| [45,55) | TPT for eligible (w/ test)_Lfx | -689 (-905 - -527) | -153 (-234 - -102) | -536 (-723 - -390) | -201 (-297 - -137) | -900 (-1,190 - -641) |
| [45,55) | TPT for eligible (w/o test)_Lfx | -835 (-1,100 - -635) | -185 (-284 - -123) | -649 (-894 - -471) | -243 (-366 - -164) | -900 (-1,190 - -641) |
| [55,65) | HHCM, no PT_None | 0 (0 - 0) | 0 (0 - 0) | 0 (0 - 0) | 0 (0 - 0) | -894 (-1,200 - -627) |
| [55,65) | TPT for eligible (w/ test)_Lfx | -521 (-665 - -408) | -116 (-171 - -77.5) | -406 (-541 - -305) | -181 (-265 - -124) | -894 (-1,200 - -627) |
| [55,65) | TPT for eligible (w/o test)_Lfx | -613 (-783 - -480) | -136 (-201 - -91.3) | -477 (-639 - -357) | -212 (-312 - -146) | -894 (-1,200 - -627) |
| [65,Inf) | HHCM, no PT_None | 0 (0 - 0) | 0 (0 - 0) | 0 (0 - 0) | 0 (0 - 0) | -1,010 (-1,360 - -689) |
| [65,Inf) | TPT for eligible (w/ test)_Lfx | -479 (-632 - -360) | -106 (-167 - -70.4) | -372 (-522 - -266) | -193 (-299 - -126) | -1,010 (-1,360 - -689) |
| [65,Inf) | TPT for eligible (w/o test)_Lfx | -542 (-702 - -419) | -121 (-186 - -80.5) | -422 (-575 - -308) | -218 (-334 - -142) | -1,010 (-1,360 - -689) |

Table A10 Resource use associated with full coverage of household contact management **cumulatively** moving up by contact age group. Tuberculosis preventive treatment (PT) is assumed to be offered to those eligible. RS = Rifampicin-susceptible; RR = Rifampicin-resistant.

| **Intervention** | **Age** | **Household contacts** | **PT courses** | **RR-ATT** | **RS-ATT** | **Cost** |
| --- | --- | --- | --- | --- | --- | --- |
| HHCM, no PT | [0,5) | 74,500 (58,600 to 92,500) | 0 (0 to 0) | 4,160 (3,130 to 5,340) | -1,080 (-1,790 to -458) | 28,700,000 (10,100,000 to 65,900,000) |
| HHCM, no PT | [5,15) | 238,000 (208,000 to 281,000) | 0 (0 to 0) | 12,500 (10,600 to 14,800) | -6,460 (-8,440 to -4,750) | 73,100,000 (35,000,000 to 136,000,000) |
| HHCM, no PT | [15,25) | 398,000 (361,000 to 440,000) | 0 (0 to 0) | 15,700 (13,800 to 18,000) | -8,500 (-10,700 to -6,650) | 91,300,000 (49,800,000 to 155,000,000) |
| HHCM, no PT | [25,35) | 509,000 (471,000 to 552,000) | 0 (0 to 0) | 18,300 (16,300 to 20,900) | -9,330 (-11,600 to -7,350) | 106,000,000 (63,400,000 to 170,000,000) |
| HHCM, no PT | [35,45) | 602,000 (564,000 to 645,000) | 0 (0 to 0) | 20,500 (18,400 to 23,200) | -9,920 (-12,300 to -7,820) | 117,000,000 (73,000,000 to 182,000,000) |
| HHCM, no PT | [45,55) | 694,000 (655,000 to 738,000) | 0 (0 to 0) | 22,500 (20,200 to 25,200) | -10,500 (-12,800 to -8,220) | 127,000,000 (81,100,000 to 195,000,000) |
| HHCM, no PT | [55,65) | 768,000 (728,000 to 814,000) | 0 (0 to 0) | 24,100 (21,700 to 27,000) | -10,900 (-13,300 to -8,660) | 135,000,000 (87,300,000 to 202,000,000) |
| HHCM, no PT | [65,Inf) | 838,000 (799,000 to 883,000) | 0 (0 to 0) | 25,600 (23,200 to 28,600) | -11,200 (-13,700 to -8,960) | 143,000,000 (94,100,000 to 211,000,000) |
| TPT for eligible (w/ test) | [0,5) | 74,500 (58,600 to 92,500) | 68,600 (54,000 to 85,300) | 3,920 (2,900 to 5,110) | -1,930 (-2,750 to -1,200) | 31,900,000 (12,800,000 to 67,700,000) |
| TPT for eligible (w/ test) | [5,15) | 238,000 (208,000 to 281,000) | 149,000 (129,000 to 174,000) | 11,800 (10,100 to 14,100) | -8,960 (-11,200 to -7,070) | 88,000,000 (50,900,000 to 148,000,000) |
| TPT for eligible (w/ test) | [15,25) | 398,000 (361,000 to 440,000) | 259,000 (234,000 to 286,000) | 14,500 (12,600 to 16,800) | -11,700 (-14,100 to -9,690) | 113,000,000 (71,600,000 to 174,000,000) |
| TPT for eligible (w/ test) | [25,35) | 509,000 (471,000 to 552,000) | 339,000 (314,000 to 367,000) | 17,000 (15,000 to 19,400) | -12,900 (-15,500 to -10,800) | 134,000,000 (89,500,000 to 198,000,000) |
| TPT for eligible (w/ test) | [35,45) | 602,000 (564,000 to 645,000) | 409,000 (383,000 to 437,000) | 18,900 (16,800 to 21,600) | -13,700 (-16,400 to -11,500) | 150,000,000 (106,000,000 to 213,000,000) |
| TPT for eligible (w/ test) | [45,55) | 694,000 (655,000 to 738,000) | 481,000 (455,000 to 510,000) | 20,700 (18,600 to 23,500) | -14,500 (-17,100 to -12,000) | 166,000,000 (122,000,000 to 232,000,000) |
| TPT for eligible (w/ test) | [55,65) | 768,000 (728,000 to 814,000) | 541,000 (515,000 to 570,000) | 22,200 (20,000 to 25,100) | -15,100 (-17,700 to -12,600) | 180,000,000 (133,000,000 to 245,000,000) |
| TPT for eligible (w/ test) | [65,Inf) | 838,000 (799,000 to 883,000) | 600,000 (573,000 to 629,000) | 23,500 (21,200 to 26,400) | -15,600 (-18,200 to -13,100) | 193,000,000 (147,000,000 to 259,000,000) |
| TPT for eligible (w/o test) | [0,5) | 74,500 (58,600 to 92,500) | 68,600 (54,000 to 85,300) | 3,920 (2,900 to 5,110) | -1,930 (-2,750 to -1,200) | 31,900,000 (12,800,000 to 67,700,000) |
| TPT for eligible (w/o test) | [5,15) | 238,000 (208,000 to 281,000) | 219,000 (192,000 to 259,000) | 11,700 (10,000 to 13,900) | -9,390 (-11,800 to -7,490) | 98,900,000 (61,000,000 to 158,000,000) |
| TPT for eligible (w/o test) | [15,25) | 398,000 (361,000 to 440,000) | 372,000 (338,000 to 412,000) | 14,200 (12,300 to 16,400) | -12,400 (-14,900 to -10,300) | 126,000,000 (82,900,000 to 188,000,000) |
| TPT for eligible (w/o test) | [25,35) | 509,000 (471,000 to 552,000) | 479,000 (444,000 to 519,000) | 16,600 (14,700 to 19,000) | -13,700 (-16,400 to -11,500) | 148,000,000 (102,000,000 to 211,000,000) |
| TPT for eligible (w/o test) | [35,45) | 602,000 (564,000 to 645,000) | 568,000 (532,000 to 609,000) | 18,500 (16,400 to 21,100) | -14,600 (-17,400 to -12,300) | 165,000,000 (120,000,000 to 226,000,000) |
| TPT for eligible (w/o test) | [45,55) | 694,000 (655,000 to 738,000) | 656,000 (619,000 to 697,000) | 20,300 (18,100 to 23,100) | -15,400 (-18,100 to -12,900) | 181,000,000 (135,000,000 to 243,000,000) |
| TPT for eligible (w/o test) | [55,65) | 768,000 (728,000 to 814,000) | 727,000 (689,000 to 770,000) | 21,700 (19,400 to 24,600) | -16,000 (-18,700 to -13,500) | 195,000,000 (148,000,000 to 259,000,000) |
| TPT for eligible (w/o test) | [65,Inf) | 838,000 (799,000 to 883,000) | 794,000 (757,000 to 836,000) | 23,000 (20,600 to 26,000) | -16,500 (-19,200 to -14,100) | 209,000,000 (161,000,000 to 275,000,000) |

Table A11 Incremental outcomes associated with full coverage of household contact management **cumulatively** moving up by contact age group. Tuberculosis preventive treatment is assumed to be offered to those eligible. RS = Rifampicin-susceptible; RR = Rifampicin-resistant.

| **Intervention** | **Age** | **Adverse Events** | **Serious Adverse Events** | **Incident TB episodes** | **Incident TB deaths** | **Prevalent TB deaths** | **DALYs** |
| --- | --- | --- | --- | --- | --- | --- | --- |
| HHCM, no PT | [0,5) | 0 (0 to 0) | 0 (0 to 0) | 0 (0 to 0) | 0 (0 to 0) | -611 (-816 to -449) | -18,000 (-24,000 to -13,200) |
| HHCM, no PT | [5,15) | 0 (0 to 0) | 0 (0 to 0) | 0 (0 to 0) | 0 (0 to 0) | -2,160 (-2,600 to -1,810) | -62,800 (-75,400 to -52,400) |
| HHCM, no PT | [15,25) | 0 (0 to 0) | 0 (0 to 0) | 0 (0 to 0) | 0 (0 to 0) | -2,880 (-3,320 to -2,490) | -82,600 (-95,400 to -71,300) |
| HHCM, no PT | [25,35) | 0 (0 to 0) | 0 (0 to 0) | 0 (0 to 0) | 0 (0 to 0) | -3,770 (-4,290 to -3,320) | -105,000 (-119,000 to -92,100) |
| HHCM, no PT | [35,45) | 0 (0 to 0) | 0 (0 to 0) | 0 (0 to 0) | 0 (0 to 0) | -4,560 (-5,120 to -4,030) | -123,000 (-138,000 to -109,000) |
| HHCM, no PT | [45,55) | 0 (0 to 0) | 0 (0 to 0) | 0 (0 to 0) | 0 (0 to 0) | -5,460 (-6,130 to -4,870) | -140,000 (-157,000 to -125,000) |
| HHCM, no PT | [55,65) | 0 (0 to 0) | 0 (0 to 0) | 0 (0 to 0) | 0 (0 to 0) | -6,350 (-7,060 to -5,670) | -153,000 (-170,000 to -137,000) |
| HHCM, no PT | [65,Inf) | 0 (0 to 0) | 0 (0 to 0) | 0 (0 to 0) | 0 (0 to 0) | -7,360 (-8,150 to -6,630) | -161,000 (-179,000 to -145,000) |
| TPT for eligible (w/ test) | [0,5) | 5,560 (4,390 to 6,950) | 2,390 (1,880 to 2,990) | -2,270 (-3,190 to -1,540) | -382 (-546 to -240) | -611 (-816 to -449) | -42,300 (-56,300 to -30,900) |
| TPT for eligible (w/ test) | [5,15) | 13,500 (11,700 to 15,900) | 5,790 (4,990 to 6,840) | -4,970 (-6,280 to -3,920) | -879 (-1,120 to -679) | -2,160 (-2,600 to -1,810) | -117,000 (-140,000 to -99,000) |
| TPT for eligible (w/ test) | [15,25) | 27,800 (25,300 to 30,500) | 11,900 (10,800 to 13,200) | -6,490 (-7,870 to -5,350) | -1,140 (-1,390 to -918) | -2,880 (-3,320 to -2,490) | -153,000 (-176,000 to -133,000) |
| TPT for eligible (w/ test) | [25,35) | 41,500 (38,600 to 44,600) | 17,800 (16,600 to 19,200) | -7,430 (-8,870 to -6,230) | -1,380 (-1,650 to -1,130) | -3,770 (-4,290 to -3,320) | -187,000 (-212,000 to -166,000) |
| TPT for eligible (w/ test) | [35,45) | 57,100 (53,700 to 60,400) | 24,500 (23,100 to 26,100) | -8,170 (-9,640 to -6,930) | -1,570 (-1,860 to -1,320) | -4,560 (-5,120 to -4,030) | -213,000 (-239,000 to -190,000) |
| TPT for eligible (w/ test) | [45,55) | 78,300 (74,400 to 82,200) | 33,600 (31,900 to 35,300) | -8,860 (-10,400 to -7,610) | -1,780 (-2,080 to -1,500) | -5,460 (-6,130 to -4,870) | -238,000 (-266,000 to -213,000) |
| TPT for eligible (w/ test) | [55,65) | 101,000 (97,200 to 106,000) | 43,500 (41,600 to 45,600) | -9,380 (-10,900 to -8,110) | -1,960 (-2,280 to -1,670) | -6,350 (-7,060 to -5,670) | -255,000 (-283,000 to -232,000) |
| TPT for eligible (w/ test) | [65,Inf) | 131,000 (126,000 to 136,000) | 56,200 (53,800 to 58,700) | -9,860 (-11,400 to -8,610) | -2,150 (-2,470 to -1,870) | -7,360 (-8,150 to -6,630) | -266,000 (-295,000 to -242,000) |
| TPT for eligible (w/o test) | [0,5) | 5,560 (4,390 to 6,950) | 2,390 (1,880 to 2,990) | -2,270 (-3,190 to -1,540) | -382 (-546 to -240) | -611 (-816 to -449) | -42,300 (-56,300 to -30,900) |
| TPT for eligible (w/o test) | [5,15) | 20,500 (17,800 to 24,500) | 8,800 (7,600 to 10,400) | -5,690 (-7,310 to -4,430) | -1,010 (-1,320 to -765) | -2,160 (-2,600 to -1,810) | -125,000 (-152,000 to -104,000) |
| TPT for eligible (w/o test) | [15,25) | 40,400 (36,700 to 44,600) | 17,400 (15,700 to 19,300) | -7,750 (-9,470 to -6,330) | -1,360 (-1,680 to -1,090) | -2,880 (-3,320 to -2,490) | -167,000 (-196,000 to -145,000) |
| TPT for eligible (w/o test) | [25,35) | 58,600 (54,500 to 63,100) | 25,200 (23,300 to 27,100) | -9,000 (-10,700 to -7,480) | -1,680 (-2,020 to -1,390) | -3,770 (-4,290 to -3,320) | -205,000 (-232,000 to -180,000) |
| TPT for eligible (w/o test) | [35,45) | 78,400 (74,000 to 83,300) | 33,700 (31,600 to 35,900) | -9,930 (-11,700 to -8,450) | -1,930 (-2,300 to -1,610) | -4,560 (-5,120 to -4,030) | -233,000 (-262,000 to -207,000) |
| TPT for eligible (w/o test) | [45,55) | 104,000 (99,300 to 110,000) | 44,800 (42,400 to 47,100) | -10,800 (-12,600 to -9,220) | -2,170 (-2,580 to -1,840) | -5,460 (-6,130 to -4,870) | -259,000 (-289,000 to -231,000) |
| TPT for eligible (w/o test) | [55,65) | 132,000 (126,000 to 138,000) | 56,500 (53,900 to 59,300) | -11,400 (-13,200 to -9,850) | -2,380 (-2,780 to -2,030) | -6,350 (-7,060 to -5,670) | -277,000 (-308,000 to -250,000) |
| TPT for eligible (w/o test) | [65,Inf) | 165,000 (159,000 to 172,000) | 71,000 (68,000 to 74,000) | -11,900 (-13,800 to -10,400) | -2,600 (-3,010 to -2,250) | -7,360 (-8,150 to -6,630) | -289,000 (-321,000 to -262,000) |

Table A12 Global and regional **total** resources and outcomes vs no HHCM in each age group.

RS = Rifampicin-susceptible; RR = Rifampicin-resistant; TB = tuberculosis; ATT = anti-TB treatment; PT = preventive treatment; DALY = disability-adjusted life-year; HHCM = household contact management

| **Region** | **Intervention** | **Household contacts** | **PT courses** | **RR-ATT** | **RS-ATT** | **Cost** | **Adverse Events** | **Serious Adverse Events** | **Incident TB episodes** | **Incident TB deaths** | **Prevalent TB deaths** | **Total TB deaths** | **DALYs** |
| --- | --- | --- | --- | --- | --- | --- | --- | --- | --- | --- | --- | --- | --- |
| AFR | HHCM, no PT | 120,000 (114,000 to 125,000) | 0 (0 to 0) | 5,960 (5,460 to 6,510) | 2,550 (2,130 to 3,030) | 9,350,000 (8,570,000 to 10,200,000) | 0 (0 to 0) | 0 (0 to 0) | 3,540 (3,130 to 3,990) | 791 (695 to 903) | 429 (388 to 478) | 1,220 (1,100 to 1,350) | 47,200 (42,100 to 52,700) |
| AFR | TPT for eligible (w/ test) | 120,000 (114,000 to 125,000) | 87,300 (83,100 to 91,500) | 5,580 (5,100 to 6,070) | 1,840 (1,560 to 2,200) | 18,300,000 (16,900,000 to 20,000,000) | 19,400 (18,500 to 20,300) | 8,310 (7,910 to 8,720) | 1,870 (1,640 to 2,160) | 420 (363 to 486) | 429 (388 to 478) | 849 (764 to 935) | 29,700 (26,500 to 33,400) |
| AFR | TPT for eligible (w/o test) | 120,000 (114,000 to 125,000) | 113,000 (108,000 to 119,000) | 5,490 (5,030 to 5,980) | 1,710 (1,430 to 2,070) | 20,400,000 (18,800,000 to 22,500,000) | 23,900 (23,000 to 25,000) | 10,300 (9,830 to 10,700) | 1,550 (1,330 to 1,850) | 350 (296 to 418) | 429 (388 to 478) | 779 (701 to 864) | 26,300 (23,100 to 30,000) |
| AMR | HHCM, no PT | 24,100 (22,800 to 25,500) | 0 (0 to 0) | 1,210 (1,110 to 1,330) | 647 (551 to 771) | 10,100,000 (8,950,000 to 11,500,000) | 0 (0 to 0) | 0 (0 to 0) | 723 (639 to 828) | 152 (132 to 175) | 86.5 (77.4 to 98.4) | 238 (214 to 268) | 9,990 (8,880 to 11,300) |
| AMR | TPT for eligible (w/ test) | 24,100 (22,800 to 25,500) | 16,800 (15,800 to 17,800) | 1,140 (1,030 to 1,250) | 451 (382 to 533) | 12,300,000 (11,100,000 to 13,700,000) | 3,690 (3,460 to 3,910) | 1,580 (1,490 to 1,680) | 392 (342 to 453) | 83.2 (71.6 to 97.2) | 86.5 (77.4 to 98.4) | 170 (153 to 190) | 6,460 (5,750 to 7,350) |
| AMR | TPT for eligible (w/o test) | 24,100 (22,800 to 25,500) | 22,900 (21,700 to 24,100) | 1,120 (1,010 to 1,230) | 405 (338 to 483) | 13,000,000 (11,700,000 to 14,500,000) | 4,790 (4,570 to 5,020) | 2,060 (1,960 to 2,160) | 317 (266 to 383) | 67.7 (56.3 to 81.9) | 86.5 (77.4 to 98.4) | 154 (138 to 174) | 5,640 (4,960 to 6,520) |
| EMR | HHCM, no PT | 27,600 (25,300 to 30,200) | 0 (0 to 0) | 1,290 (1,120 to 1,510) | 683 (530 to 880) | 1,780,000 (1,590,000 to 2,010,000) | 0 (0 to 0) | 0 (0 to 0) | 691 (559 to 856) | 155 (125 to 192) | 98.7 (81.8 to 120) | 254 (214 to 306) | 9,870 (8,150 to 12,100) |
| EMR | TPT for eligible (w/ test) | 27,600 (25,300 to 30,200) | 20,000 (18,300 to 21,700) | 1,250 (1,080 to 1,470) | 520 (396 to 677) | 3,150,000 (2,810,000 to 3,570,000) | 4,490 (4,130 to 4,880) | 1,930 (1,760 to 2,110) | 434 (343 to 544) | 100 (79.1 to 126) | 98.7 (81.8 to 120) | 199 (166 to 239) | 7,140 (5,830 to 8,790) |
| EMR | TPT for eligible (w/o test) | 27,600 (25,300 to 30,200) | 26,200 (24,000 to 28,700) | 1,240 (1,070 to 1,450) | 486 (364 to 637) | 3,560,000 (3,130,000 to 4,090,000) | 5,530 (5,120 to 5,960) | 2,370 (2,180 to 2,580) | 379 (290 to 500) | 88.7 (67.8 to 119) | 98.7 (81.8 to 120) | 187 (156 to 230) | 6,540 (5,260 to 8,310) |
| EUR | HHCM, no PT | 176,000 (165,000 to 189,000) | 0 (0 to 0) | 9,580 (8,350 to 10,900) | 3,580 (2,810 to 4,580) | 211,000,000 (140,000,000 to 309,000,000) | 0 (0 to 0) | 0 (0 to 0) | 6,510 (5,660 to 7,520) | 1,440 (1,210 to 1,700) | 646 (560 to 748) | 2,080 (1,820 to 2,390) | 85,500 (74,300 to 98,200) |
| EUR | TPT for eligible (w/ test) | 176,000 (165,000 to 189,000) | 122,000 (115,000 to 132,000) | 8,890 (7,810 to 10,000) | 2,760 (2,160 to 3,500) | 216,000,000 (151,000,000 to 305,000,000) | 25,400 (23,900 to 27,200) | 10,900 (10,200 to 11,800) | 3,910 (3,380 to 4,570) | 880 (735 to 1,040) | 646 (560 to 748) | 1,530 (1,330 to 1,740) | 58,100 (50,500 to 66,900) |
| EUR | TPT for eligible (w/o test) | 176,000 (165,000 to 189,000) | 167,000 (156,000 to 179,000) | 8,730 (7,650 to 9,880) | 2,580 (2,010 to 3,270) | 220,000,000 (156,000,000 to 307,000,000) | 34,000 (32,100 to 36,100) | 14,600 (13,800 to 15,500) | 3,340 (2,800 to 4,040) | 750 (613 to 923) | 646 (560 to 748) | 1,400 (1,210 to 1,610) | 51,900 (44,700 to 60,900) |
| SEA | HHCM, no PT | 382,000 (345,000 to 421,000) | 0 (0 to 0) | 18,800 (15,700 to 22,500) | 8,100 (5,840 to 10,900) | 36,200,000 (28,200,000 to 45,900,000) | 0 (0 to 0) | 0 (0 to 0) | 10,300 (8,130 to 12,900) | 2,280 (1,820 to 2,840) | 1,370 (1,100 to 1,680) | 3,640 (3,010 to 4,430) | 144,000 (116,000 to 178,000) |
| SEA | TPT for eligible (w/ test) | 382,000 (345,000 to 421,000) | 275,000 (250,000 to 300,000) | 18,100 (15,100 to 21,500) | 6,370 (4,500 to 8,710) | 56,900,000 (47,900,000 to 68,700,000) | 60,700 (55,900 to 66,100) | 26,000 (23,900 to 28,400) | 6,600 (5,140 to 8,450) | 1,490 (1,150 to 1,920) | 1,370 (1,100 to 1,680) | 2,860 (2,330 to 3,500) | 105,000 (84,200 to 130,000) |
| SEA | TPT for eligible (w/o test) | 382,000 (345,000 to 421,000) | 362,000 (327,000 to 399,000) | 17,900 (15,000 to 21,300) | 5,970 (4,200 to 8,280) | 62,900,000 (52,400,000 to 76,900,000) | 75,800 (69,700 to 82,400) | 32,500 (29,800 to 35,400) | 5,820 (4,360 to 7,660) | 1,330 (995 to 1,780) | 1,370 (1,100 to 1,680) | 2,700 (2,190 to 3,340) | 96,700 (75,800 to 122,000) |
| WPR | HHCM, no PT | 109,000 (97,400 to 124,000) | 0 (0 to 0) | 5,040 (4,250 to 6,150) | 2,530 (1,940 to 3,280) | 24,100,000 (15,100,000 to 42,000,000) | 0 (0 to 0) | 0 (0 to 0) | 2,810 (2,070 to 3,880) | 661 (493 to 903) | 395 (317 to 492) | 1,060 (830 to 1,360) | 41,800 (31,600 to 56,400) |
| WPR | TPT for eligible (w/ test) | 109,000 (97,400 to 124,000) | 79,400 (70,200 to 92,300) | 4,900 (4,120 to 6,000) | 1,820 (1,380 to 2,350) | 36,400,000 (26,500,000 to 55,200,000) | 17,400 (16,100 to 18,900) | 7,480 (6,860 to 8,190) | 1,460 (1,110 to 2,020) | 346 (262 to 474) | 395 (317 to 492) | 741 (597 to 939) | 26,700 (20,900 to 35,200) |
| WPR | TPT for eligible (w/o test) | 109,000 (97,400 to 124,000) | 103,000 (92,500 to 117,000) | 4,860 (4,080 to 5,960) | 1,640 (1,230 to 2,140) | 39,500,000 (29,200,000 to 58,200,000) | 21,500 (20,000 to 22,900) | 9,200 (8,560 to 9,950) | 1,200 (842 to 1,770) | 285 (201 to 418) | 395 (317 to 492) | 680 (541 to 867) | 23,600 (18,000 to 32,000) |
| Global | HHCM, no PT | 838,000 (799,000 to 883,000) | 0 (0 to 0) | 41,900 (38,400 to 45,900) | 18,100 (15,500 to 21,100) | 293,000,000 (221,000,000 to 392,000,000) | 0 (0 to 0) | 0 (0 to 0) | 24,500 (21,900 to 27,600) | 5,470 (4,870 to 6,180) | 3,020 (2,730 to 3,390) | 8,490 (7,700 to 9,380) | 338,000 (305,000 to 378,000) |
| Global | TPT for eligible (w/ test) | 838,000 (799,000 to 883,000) | 600,000 (573,000 to 629,000) | 39,800 (36,600 to 43,600) | 13,800 (11,700 to 16,300) | 343,000,000 (277,000,000 to 434,000,000) | 131,000 (126,000 to 136,000) | 56,200 (53,800 to 58,700) | 14,700 (12,900 to 16,600) | 3,320 (2,920 to 3,810) | 3,020 (2,730 to 3,390) | 6,340 (5,760 to 7,030) | 233,000 (209,000 to 261,000) |
| Global | TPT for eligible (w/o test) | 838,000 (799,000 to 883,000) | 794,000 (757,000 to 836,000) | 39,300 (36,100 to 43,200) | 12,800 (10,900 to 15,200) | 359,000,000 (294,000,000 to 449,000,000) | 165,000 (159,000 to 172,000) | 71,000 (68,000 to 74,000) | 12,600 (11,000 to 14,600) | 2,870 (2,490 to 3,390) | 3,020 (2,730 to 3,390) | 5,900 (5,330 to 6,570) | 211,000 (188,000 to 238,000) |

Table A13 Global and regional **incremental** resources and outcomes vs no HHCM in each age group

RS = Rifampicin-susceptible; RR = Rifampicin-resistant; TB = tuberculosis; ATT = anti-TB treatment; PT = preventive treatment; DALY = disability-adjusted life-year; HHCM = household contact management

| **Region** | **Intervention** | **Household contacts** | **PT courses** | **RR-ATT** | **RS-ATT** | **Cost** | **Adverse Events** | **Serious Adverse Events** | **Incident TB episodes** | **Incident TB deaths** | **Prevalent TB deaths** | **Total TB deaths** | **DALYs** |
| --- | --- | --- | --- | --- | --- | --- | --- | --- | --- | --- | --- | --- | --- |
| AFR | HHCM, no PT | 120,000 (114,000 to 125,000) | 0 (0 to 0) | 3,610 (3,240 to 4,040) | -1,490 (-1,910 to -1,050) | 4,790,000 (4,230,000 to 5,390,000) | 0 (0 to 0) | 0 (0 to 0) | 0 (0 to 0) | 0 (0 to 0) | -1,050 (-1,170 to -930) | -1,050 (-1,170 to -930) | -21,600 (-24,200 to -19,200) |
| AFR | TPT for eligible (w/ test) | 120,000 (114,000 to 125,000) | 87,300 (83,100 to 91,500) | 3,220 (2,850 to 3,630) | -2,190 (-2,650 to -1,690) | 13,800,000 (12,400,000 to 15,300,000) | 19,400 (18,500 to 20,300) | 8,310 (7,910 to 8,720) | -1,670 (-1,930 to -1,440) | -371 (-433 to -318) | -1,050 (-1,170 to -930) | -1,420 (-1,570 to -1,280) | -39,100 (-43,400 to -35,200) |
| AFR | TPT for eligible (w/o test) | 120,000 (114,000 to 125,000) | 113,000 (108,000 to 119,000) | 3,140 (2,760 to 3,560) | -2,320 (-2,820 to -1,820) | 15,900,000 (14,300,000 to 17,800,000) | 23,900 (23,000 to 25,000) | 10,300 (9,830 to 10,700) | -1,990 (-2,340 to -1,710) | -441 (-528 to -373) | -1,050 (-1,170 to -930) | -1,490 (-1,650 to -1,340) | -42,500 (-47,600 to -37,900) |
| AMR | HHCM, no PT | 24,100 (22,800 to 25,500) | 0 (0 to 0) | 707 (625 to 798) | -512 (-621 to -420) | 3,910,000 (3,140,000 to 4,760,000) | 0 (0 to 0) | 0 (0 to 0) | 0 (0 to 0) | 0 (0 to 0) | -186 (-208 to -165) | -186 (-208 to -165) | -4,390 (-4,930 to -3,860) |
| AMR | TPT for eligible (w/ test) | 24,100 (22,800 to 25,500) | 16,800 (15,800 to 17,800) | 629 (548 to 720) | -709 (-844 to -600) | 6,090,000 (5,240,000 to 7,090,000) | 3,690 (3,460 to 3,910) | 1,580 (1,490 to 1,680) | -331 (-392 to -280) | -68.4 (-81.5 to -57.4) | -186 (-208 to -165) | -254 (-281 to -229) | -7,920 (-8,880 to -7,070) |
| AMR | TPT for eligible (w/o test) | 24,100 (22,800 to 25,500) | 22,900 (21,700 to 24,100) | 609 (528 to 700) | -755 (-893 to -639) | 6,800,000 (5,840,000 to 7,900,000) | 4,790 (4,570 to 5,020) | 2,060 (1,960 to 2,160) | -406 (-482 to -345) | -83.9 (-100 to -70.5) | -186 (-208 to -165) | -270 (-299 to -242) | -8,740 (-9,790 to -7,750) |
| EMR | HHCM, no PT | 27,600 (25,300 to 30,200) | 0 (0 to 0) | 906 (773 to 1,080) | -618 (-816 to -432) | 895,000 (760,000 to 1,070,000) | 0 (0 to 0) | 0 (0 to 0) | 0 (0 to 0) | 0 (0 to 0) | -249 (-291 to -212) | -249 (-291 to -212) | -5,300 (-6,290 to -4,430) |
| EMR | TPT for eligible (w/ test) | 27,600 (25,300 to 30,200) | 20,000 (18,300 to 21,700) | 867 (735 to 1,040) | -782 (-993 to -588) | 2,260,000 (1,960,000 to 2,610,000) | 4,490 (4,130 to 4,880) | 1,930 (1,760 to 2,110) | -257 (-331 to -199) | -55 (-71.6 to -41.7) | -249 (-291 to -212) | -304 (-353 to -263) | -8,030 (-9,520 to -6,850) |
| EMR | TPT for eligible (w/o test) | 27,600 (25,300 to 30,200) | 26,200 (24,000 to 28,700) | 856 (724 to 1,030) | -815 (-1,030 to -610) | 2,670,000 (2,280,000 to 3,150,000) | 5,530 (5,120 to 5,960) | 2,370 (2,180 to 2,580) | -312 (-413 to -237) | -66.3 (-89 to -49.2) | -249 (-291 to -212) | -316 (-366 to -273) | -8,620 (-10,400 to -7,310) |
| EUR | HHCM, no PT | 176,000 (165,000 to 189,000) | 0 (0 to 0) | 5,080 (4,010 to 6,140) | -1,190 (-2,020 to -395) | 102,000,000 (56,500,000 to 168,000,000) | 0 (0 to 0) | 0 (0 to 0) | 0 (0 to 0) | 0 (0 to 0) | -1,440 (-1,730 to -1,170) | -1,440 (-1,730 to -1,170) | -33,000 (-39,700 to -26,800) |
| EUR | TPT for eligible (w/ test) | 176,000 (165,000 to 189,000) | 122,000 (115,000 to 132,000) | 4,390 (3,300 to 5,480) | -2,010 (-2,920 to -1,190) | 107,000,000 (62,100,000 to 169,000,000) | 25,400 (23,900 to 27,200) | 10,900 (10,200 to 11,800) | -2,610 (-3,220 to -2,140) | -559 (-695 to -439) | -1,440 (-1,730 to -1,170) | -2,000 (-2,360 to -1,690) | -60,400 (-71,300 to -51,200) |
| EUR | TPT for eligible (w/o test) | 176,000 (165,000 to 189,000) | 167,000 (156,000 to 179,000) | 4,230 (3,080 to 5,320) | -2,190 (-3,170 to -1,320) | 110,000,000 (65,600,000 to 171,000,000) | 34,000 (32,100 to 36,100) | 14,600 (13,800 to 15,500) | -3,170 (-3,920 to -2,600) | -688 (-858 to -542) | -1,440 (-1,730 to -1,170) | -2,130 (-2,510 to -1,790) | -66,600 (-78,700 to -56,300) |
| SEA | HHCM, no PT | 382,000 (345,000 to 421,000) | 0 (0 to 0) | 11,400 (9,260 to 13,900) | -5,260 (-7,440 to -3,260) | 18,000,000 (13,300,000 to 23,600,000) | 0 (0 to 0) | 0 (0 to 0) | 0 (0 to 0) | 0 (0 to 0) | -3,380 (-4,080 to -2,760) | -3,380 (-4,080 to -2,760) | -72,600 (-87,100 to -59,700) |
| SEA | TPT for eligible (w/ test) | 382,000 (345,000 to 421,000) | 275,000 (250,000 to 300,000) | 10,600 (8,610 to 13,100) | -6,990 (-9,320 to -4,990) | 38,800,000 (32,400,000 to 46,700,000) | 60,700 (55,900 to 66,100) | 26,000 (23,900 to 28,400) | -3,650 (-4,850 to -2,700) | -781 (-1,050 to -569) | -3,380 (-4,080 to -2,760) | -4,160 (-4,930 to -3,450) | -111,000 (-133,000 to -92,200) |
| SEA | TPT for eligible (w/o test) | 382,000 (345,000 to 421,000) | 362,000 (327,000 to 399,000) | 10,500 (8,440 to 13,000) | -7,390 (-9,820 to -5,320) | 44,800,000 (37,000,000 to 54,900,000) | 75,800 (69,700 to 82,400) | 32,500 (29,800 to 35,400) | -4,430 (-6,090 to -3,180) | -944 (-1,280 to -663) | -3,380 (-4,080 to -2,760) | -4,320 (-5,140 to -3,580) | -120,000 (-143,000 to -99,300) |
| WPR | HHCM, no PT | 109,000 (97,400 to 124,000) | 0 (0 to 0) | 3,910 (3,180 to 4,970) | -2,160 (-2,880 to -1,480) | 13,400,000 (6,020,000 to 30,100,000) | 0 (0 to 0) | 0 (0 to 0) | 0 (0 to 0) | 0 (0 to 0) | -1,060 (-1,240 to -901) | -1,060 (-1,240 to -901) | -24,500 (-30,200 to -20,600) |
| WPR | TPT for eligible (w/ test) | 109,000 (97,400 to 124,000) | 79,400 (70,200 to 92,300) | 3,760 (3,040 to 4,820) | -2,880 (-3,730 to -2,160) | 25,700,000 (17,300,000 to 43,600,000) | 17,400 (16,100 to 18,900) | 7,480 (6,860 to 8,190) | -1,350 (-1,950 to -936) | -315 (-442 to -216) | -1,060 (-1,240 to -901) | -1,370 (-1,660 to -1,150) | -39,700 (-51,100 to -31,800) |
| WPR | TPT for eligible (w/o test) | 109,000 (97,400 to 124,000) | 103,000 (92,500 to 117,000) | 3,730 (3,000 to 4,790) | -3,060 (-3,940 to -2,270) | 28,800,000 (20,100,000 to 46,300,000) | 21,500 (20,000 to 22,900) | 9,200 (8,560 to 9,950) | -1,610 (-2,310 to -1,110) | -376 (-527 to -262) | -1,060 (-1,240 to -901) | -1,430 (-1,730 to -1,200) | -42,700 (-55,000 to -34,100) |
| Global | HHCM, no PT | 838,000 (799,000 to 883,000) | 0 (0 to 0) | 25,600 (23,200 to 28,600) | -11,200 (-13,700 to -8,960) | 143,000,000 (94,100,000 to 211,000,000) | 0 (0 to 0) | 0 (0 to 0) | 0 (0 to 0) | 0 (0 to 0) | -7,360 (-8,150 to -6,630) | -7,360 (-8,150 to -6,630) | -161,000 (-179,000 to -145,000) |
| Global | TPT for eligible (w/ test) | 838,000 (799,000 to 883,000) | 600,000 (573,000 to 629,000) | 23,500 (21,200 to 26,400) | -15,600 (-18,200 to -13,100) | 193,000,000 (147,000,000 to 259,000,000) | 131,000 (126,000 to 136,000) | 56,200 (53,800 to 58,700) | -9,860 (-11,400 to -8,610) | -2,150 (-2,470 to -1,870) | -7,360 (-8,150 to -6,630) | -9,510 (-10,400 to -8,680) | -266,000 (-295,000 to -242,000) |
| Global | TPT for eligible (w/o test) | 838,000 (799,000 to 883,000) | 794,000 (757,000 to 836,000) | 23,000 (20,600 to 26,000) | -16,500 (-19,200 to -14,100) | 209,000,000 (161,000,000 to 275,000,000) | 165,000 (159,000 to 172,000) | 71,000 (68,000 to 74,000) | -11,900 (-13,800 to -10,400) | -2,600 (-3,010 to -2,250) | -7,360 (-8,150 to -6,630) | -9,960 (-10,900 to -9,100) | -289,000 (-321,000 to -262,000) |

Table A14 Percentage of global incremental costs and DALYs vs no HHCM in each region

PT = preventive treatment; DALY = disability-adjusted life-year; HHCM = household contact management; AFR = African Region; AMR = American Region; EUR = European Region; EMR = Eastern Mediterranean Region; SEA = South-East Asian Region; WPR = Western Pacific Region

| **Intervention** | **Region** | **Cost** | **DALYs** |
| --- | --- | --- | --- |
| TPT for eligible (w/ test) | AFR | 7.26 (5.33 to 9.58) | 14.7 (12.8 to 16.8) |
| TPT for eligible (w/ test) | AMR | 3.21 (2.34 to 4.25) | 2.98 (2.58 to 3.43) |
| TPT for eligible (w/ test) | EMR | 1.19 (0.848 to 1.59) | 3.02 (2.49 to 3.64) |
| TPT for eligible (w/ test) | EUR | 54.5 (41.2 to 66.5) | 22.7 (19.2 to 26.1) |
| TPT for eligible (w/ test) | SEA | 20.4 (14.1 to 27.3) | 41.7 (36.7 to 46.8) |
| TPT for eligible (w/ test) | WPR | 13.4 (8.54 to 21.5) | 14.9 (12.1 to 18.6) |
| TPT for eligible (w/o test) | AFR | 7.73 (5.74 to 10.1) | 14.7 (12.7 to 16.9) |
| TPT for eligible (w/o test) | AMR | 3.31 (2.44 to 4.35) | 3.03 (2.62 to 3.5) |
| TPT for eligible (w/o test) | EMR | 1.3 (0.935 to 1.74) | 2.99 (2.46 to 3.69) |
| TPT for eligible (w/o test) | EUR | 52 (39 to 63.7) | 23.1 (19.6 to 26.6) |
| TPT for eligible (w/o test) | SEA | 21.8 (15.4 to 28.9) | 41.4 (36.4 to 46.6) |
| TPT for eligible (w/o test) | WPR | 13.9 (9.18 to 21.5) | 14.8 (11.9 to 18.4) |

Table A15 Percentage of resources and outcomes across all ages if using PT with testing vs no testing, regionally and globally

RS = Rifampicin-susceptible; RR = Rifampicin-resistant; TB = tuberculosis; ATT = anti-TB treatment; PT = preventive treatment; DALY = disability-adjusted life-year; HHCM = household contact management; AFR = African Region; AMR = American Region; EUR = European Region; EMR = Eastern Mediterranean Region; SEA = South-East Asian Region; WPR = Western Pacific Region

| **Region** | **Metric** | **PT courses** | **RR-ATT** | **RS-ATT** | **Cost** | **Adverse Events** | **Serious Adverse Events** | **Incident TB episodes** | **Incident TB deaths** | **Total TB deaths** | **DALYs** |
| --- | --- | --- | --- | --- | --- | --- | --- | --- | --- | --- | --- |
| AFR | incremental | 77 (75.4 to 78.4) | 103 (101 to 104) | 94.3 (89.9 to 98.9) | 86.7 (84.2 to 88.6) | 80.9 (79 to 82.8) | 80.9 (78.9 to 82.8) | 84.1 (78.7 to 90.9) | 84.4 (78.9 to 91) | 95.4 (93.2 to 97.4) | 92 (88.5 to 95.8) |
| AFR | total | 77 (75.4 to 78.4) | 102 (101 to 102) | 108 (101 to 115) | 89.6 (87.6 to 91.3) | 80.9 (79 to 82.8) | 80.9 (78.9 to 82.8) | 121 (110 to 133) | 120 (110 to 132) | 109 (105 to 114) | 113 (106 to 120) |
| AMR | incremental | 73.3 (71.3 to 75.1) | 103 (102 to 105) | 93.9 (90.9 to 96.9) | 89.6 (87.1 to 92.2) | 77 (74.5 to 79.4) | 77 (74.4 to 79.4) | 81.7 (75.4 to 88.7) | 81.7 (75.1 to 89.2) | 94.3 (91.6 to 96.9) | 90.7 (86.8 to 94.7) |
| AMR | total | 73.3 (71.3 to 75.1) | 102 (101 to 103) | 112 (105 to 119) | 94.6 (93.2 to 96) | 77 (74.5 to 79.4) | 77 (74.4 to 79.4) | 124 (113 to 138) | 123 (111 to 138) | 110 (105 to 116) | 115 (107 to 123) |
| EMR | incremental | 76.3 (74 to 78.3) | 101 (101 to 102) | 95.9 (91.4 to 101) | 84.8 (81 to 87.9) | 81.1 (78.9 to 83.3) | 81.1 (78.8 to 83.3) | 82.8 (73.2 to 97.8) | 83.2 (73 to 99.4) | 96.4 (93.5 to 99.9) | 93.2 (88.1 to 99.8) |
| EMR | total | 76.3 (74 to 78.3) | 101 (100 to 101) | 107 (99.2 to 116) | 88.6 (85.2 to 91.1) | 81.1 (78.9 to 83.3) | 81.1 (78.8 to 83.3) | 115 (101 to 129) | 113 (100 to 127) | 106 (100 to 112) | 109 (100 to 118) |
| EUR | incremental | 73.3 (71.5 to 75.1) | 104 (101 to 108) | 91.7 (84.3 to 96.6) | 96.7 (92.1 to 103) | 74.8 (72.7 to 77.2) | 74.8 (72.7 to 77.1) | 82.2 (76.2 to 89.7) | 81.3 (75.2 to 88.9) | 93.9 (91 to 96.5) | 90.8 (86.7 to 95.2) |
| EUR | total | 73.3 (71.5 to 75.1) | 102 (101 to 104) | 107 (103 to 112) | 98.3 (96.1 to 101) | 74.8 (72.7 to 77.2) | 74.8 (72.7 to 77.1) | 117 (108 to 127) | 117 (109 to 127) | 109 (105 to 114) | 112 (106 to 119) |
| SEA | incremental | 75.9 (73.3 to 78) | 102 (99.9 to 104) | 94.6 (88.8 to 100) | 86.6 (82.5 to 90.2) | 80 (77.8 to 82) | 80 (77.9 to 82) | 82.7 (71.6 to 97) | 83.1 (73.1 to 95.9) | 96.3 (93.4 to 99.3) | 93.1 (87.3 to 99.1) |
| SEA | total | 75.9 (73.3 to 78) | 101 (99.9 to 102) | 107 (99.8 to 115) | 90.5 (87.3 to 93.2) | 80 (77.8 to 82) | 80 (77.9 to 82) | 114 (102 to 129) | 113 (102 to 125) | 106 (101 to 112) | 109 (101 to 118) |
| WPR | incremental | 76.8 (74 to 80.1) | 101 (101 to 102) | 94.3 (88.9 to 99.8) | 88.8 (83.6 to 94) | 81.3 (78.5 to 83.9) | 81.3 (78.5 to 83.9) | 83.6 (73 to 95.7) | 83.9 (73.4 to 96.3) | 95.8 (92.5 to 99.1) | 92.9 (87.4 to 98.5) |
| WPR | total | 76.8 (74 to 80.1) | 101 (100 to 101) | 111 (100 to 124) | 91.9 (88.2 to 95.2) | 81.3 (78.5 to 83.9) | 81.3 (78.5 to 83.9) | 123 (105 to 148) | 123 (104 to 147) | 109 (102 to 118) | 113 (102 to 127) |
| Global | incremental | 75.6 (74.3 to 76.7) | 102 (101 to 104) | 94.2 (91.1 to 96.9) | 92.3 (89.2 to 95.6) | 79.2 (78 to 80.3) | 79.2 (78 to 80.3) | 82.8 (77.8 to 88.5) | 82.8 (78.3 to 88.1) | 95.5 (94 to 97) | 92.3 (89.5 to 95) |
| Global | total | 75.6 (74.3 to 76.7) | 101 (101 to 102) | 108 (104 to 112) | 95.5 (93.7 to 97.5) | 79.2 (78 to 80.3) | 79.2 (78 to 80.3) | 116 (110 to 124) | 116 (110 to 122) | 108 (105 to 111) | 111 (106 to 116) |

Table A16 Regional and global ICERs for HHCM vs no HHCM in each age group

ICER = incremental cost-effectiveness ratio ($/DALY); DALY = disability-adjusted life-year; HHCM = household contact management; TPT = tuberculosis preventive treatment; AFR = African Region; AMR = American Region; EUR = European Region; EMR = Eastern Mediterranean Region; SEA = South-East Asian Region; WPR = Western Pacific Region

| **Intervention** | **Region** | **[0,5)** | **[5,15)** | **[15,25)** | **[25,35)** | **[35,45)** | **[45,55)** | **[55,65)** | **[65,Inf)** |
| --- | --- | --- | --- | --- | --- | --- | --- | --- | --- |
| TPT for eligible (w/ test) | AFR | 250 | 309 | 354 | 311 | 362 | 416 | 490 | 860 |
| TPT for eligible (w/ test) | AMR | 671 | 684 | 748 | 746 | 774 | 830 | 920 | 1,400 |
| TPT for eligible (w/ test) | EMR | 228 | 312 | 254 | 225 | 241 | 275 | 331 | 486 |
| TPT for eligible (w/ test) | EUR | 1,750 | 2,070 | 1,530 | 1,500 | 1,540 | 1,560 | 1,760 | 2,680 |
| TPT for eligible (w/ test) | SEA | 276 | 399 | 338 | 276 | 300 | 332 | 381 | 598 |
| TPT for eligible (w/ test) | WPR | 618 | 557 | 656 | 571 | 630 | 712 | 814 | 1,300 |
| TPT for eligible (w/ test) | Global | 754 | 749 | 704 | 609 | 622 | 659 | 765 | 1,200 |
| TPT for eligible (w/o test) | AFR | 250 | 389 | 337 | 307 | 358 | 413 | 490 | 868 |
| TPT for eligible (w/o test) | AMR | 671 | 754 | 695 | 720 | 763 | 828 | 927 | 1,410 |
| TPT for eligible (w/o test) | EMR | 228 | 401 | 248 | 225 | 242 | 275 | 330 | 483 |
| TPT for eligible (w/o test) | EUR | 1,750 | 2,010 | 1,300 | 1,340 | 1,390 | 1,450 | 1,660 | 2,600 |
| TPT for eligible (w/o test) | SEA | 276 | 497 | 323 | 273 | 300 | 332 | 381 | 594 |
| TPT for eligible (w/o test) | WPR | 618 | 641 | 645 | 582 | 645 | 728 | 831 | 1,310 |
| TPT for eligible (w/o test) | Global | 754 | 810 | 639 | 580 | 602 | 647 | 757 | 1,200 |

Table A17 Estimated numbers of musculoskeletal adverse events and grade 2+ musculoskeletal adverse events associated with providing full TPT coverage in each age group, by strategy and region

TPT = tuberculosis preventive treatment; AFR = African Region; AMR = American Region; EUR = European Region; EMR = Eastern Mediterranean Region; SEA = South-East Asian Region; WPR = Western Pacific Region

| **WHO region** | **Intervention** | **Type** | **[0,5)** | **[5,15)** | **[15,25)** | **[25,35)** | **[35,45)** | **[45,55)** | **[55,65)** | **[65,Inf)** |
| --- | --- | --- | --- | --- | --- | --- | --- | --- | --- | --- |
| AFR | TPT for eligible (w/ test) | Adverse Events | 785 (656 - 949) | 1,160 (956 - 1,390) | 2,160 (1,920 - 2,430) | 2,050 (1,850 - 2,270) | 2,360 (2,150 - 2,620) | 3,170 (2,840 - 3,550) | 3,440 (3,050 - 3,870) | 4,370 (3,830 - 4,950) |
| AFR | TPT for eligible (w/ test) | Serious Adverse Events | 336 (281 - 405) | 498 (408 - 602) | 926 (817 - 1,050) | 879 (784 - 988) | 1,010 (913 - 1,130) | 1,360 (1,210 - 1,530) | 1,480 (1,300 - 1,650) | 1,880 (1,640 - 2,130) |
| AFR | TPT for eligible (w/o test) | Adverse Events | 785 (656 - 949) | 2,160 (1,820 - 2,550) | 2,900 (2,580 - 3,270) | 2,630 (2,390 - 2,910) | 2,880 (2,640 - 3,160) | 3,750 (3,400 - 4,140) | 3,970 (3,580 - 4,370) | 4,990 (4,520 - 5,530) |
| AFR | TPT for eligible (w/o test) | Serious Adverse Events | 336 (281 - 405) | 925 (776 - 1,110) | 1,250 (1,100 - 1,400) | 1,130 (1,010 - 1,260) | 1,240 (1,120 - 1,360) | 1,610 (1,440 - 1,780) | 1,700 (1,540 - 1,880) | 2,140 (1,920 - 2,400) |
| AMR | TPT for eligible (w/ test) | Adverse Events | 165 (136 - 199) | 228 (185 - 279) | 396 (348 - 455) | 371 (332 - 414) | 428 (382 - 487) | 597 (520 - 683) | 654 (572 - 749) | 862 (721 - 999) |
| AMR | TPT for eligible (w/ test) | Serious Adverse Events | 70.9 (58.4 - 86.7) | 97.6 (79.8 - 121) | 170 (146 - 195) | 160 (142 - 180) | 184 (163 - 212) | 257 (224 - 295) | 280 (243 - 326) | 371 (311 - 432) |
| AMR | TPT for eligible (w/o test) | Adverse Events | 165 (136 - 199) | 431 (356 - 522) | 579 (510 - 662) | 523 (472 - 581) | 567 (514 - 629) | 749 (673 - 842) | 787 (704 - 883) | 1,000 (882 - 1,140) |
| AMR | TPT for eligible (w/o test) | Serious Adverse Events | 70.9 (58.4 - 86.7) | 185 (152 - 226) | 248 (215 - 284) | 225 (200 - 253) | 244 (219 - 274) | 322 (287 - 363) | 337 (300 - 382) | 431 (376 - 492) |
| EMR | TPT for eligible (w/ test) | Adverse Events | 192 (133 - 272) | 267 (191 - 370) | 478 (382 - 595) | 463 (388 - 557) | 543 (448 - 660) | 734 (612 - 887) | 817 (664 - 1,010) | 1,040 (839 - 1,270) |
| EMR | TPT for eligible (w/ test) | Serious Adverse Events | 82.3 (58.4 - 119) | 115 (82.1 - 159) | 204 (162 - 259) | 198 (163 - 243) | 233 (189 - 286) | 315 (257 - 383) | 352 (283 - 446) | 447 (355 - 551) |
| EMR | TPT for eligible (w/o test) | Adverse Events | 192 (133 - 272) | 504 (366 - 691) | 667 (537 - 825) | 611 (510 - 732) | 675 (564 - 820) | 865 (728 - 1,040) | 924 (764 - 1,130) | 1,140 (933 - 1,360) |
| EMR | TPT for eligible (w/o test) | Serious Adverse Events | 82.3 (58.4 - 119) | 216 (156 - 297) | 285 (227 - 357) | 262 (218 - 317) | 290 (236 - 354) | 372 (307 - 449) | 397 (322 - 500) | 490 (393 - 594) |
| EUR | TPT for eligible (w/ test) | Adverse Events | 1,160 (900 - 1,480) | 1,590 (1,210 - 2,070) | 2,810 (2,350 - 3,320) | 2,620 (2,310 - 2,990) | 2,910 (2,540 - 3,360) | 3,940 (3,380 - 4,550) | 4,310 (3,710 - 5,020) | 5,590 (4,660 - 6,710) |
| EUR | TPT for eligible (w/ test) | Serious Adverse Events | 498 (383 - 648) | 682 (523 - 893) | 1,210 (999 - 1,450) | 1,120 (980 - 1,280) | 1,250 (1,070 - 1,440) | 1,690 (1,450 - 1,980) | 1,860 (1,600 - 2,180) | 2,400 (1,990 - 2,920) |
| EUR | TPT for eligible (w/o test) | Adverse Events | 1,160 (900 - 1,480) | 2,980 (2,300 - 3,840) | 4,030 (3,370 - 4,790) | 3,680 (3,240 - 4,210) | 3,940 (3,460 - 4,520) | 5,200 (4,550 - 5,900) | 5,550 (4,840 - 6,360) | 6,930 (5,990 - 7,980) |
| EUR | TPT for eligible (w/o test) | Serious Adverse Events | 498 (383 - 648) | 1,280 (974 - 1,650) | 1,730 (1,440 - 2,070) | 1,580 (1,380 - 1,800) | 1,690 (1,470 - 1,950) | 2,230 (1,920 - 2,580) | 2,390 (2,060 - 2,760) | 2,980 (2,540 - 3,460) |
| SEA | TPT for eligible (w/ test) | Adverse Events | 2,640 (1,850 - 3,710) | 3,710 (2,560 - 5,580) | 6,640 (5,200 - 8,360) | 6,400 (5,170 - 7,690) | 7,300 (6,060 - 8,600) | 9,920 (8,030 - 12,000) | 10,700 (8,710 - 13,100) | 13,800 (11,100 - 17,300) |
| SEA | TPT for eligible (w/ test) | Serious Adverse Events | 1,130 (782 - 1,630) | 1,590 (1,100 - 2,340) | 2,860 (2,220 - 3,680) | 2,750 (2,220 - 3,370) | 3,140 (2,570 - 3,840) | 4,240 (3,400 - 5,250) | 4,590 (3,710 - 5,520) | 5,930 (4,710 - 7,440) |
| SEA | TPT for eligible (w/o test) | Adverse Events | 2,640 (1,850 - 3,710) | 6,980 (4,810 - 10,400) | 9,190 (7,180 - 11,600) | 8,380 (6,740 - 10,000) | 9,180 (7,640 - 10,900) | 12,000 (9,700 - 14,600) | 12,500 (10,200 - 15,200) | 15,400 (12,600 - 18,800) |
| SEA | TPT for eligible (w/o test) | Serious Adverse Events | 1,130 (782 - 1,630) | 2,990 (2,070 - 4,380) | 3,960 (3,050 - 5,060) | 3,600 (2,910 - 4,400) | 3,950 (3,260 - 4,850) | 5,130 (4,100 - 6,330) | 5,360 (4,300 - 6,490) | 6,610 (5,320 - 8,170) |
| WPR | TPT for eligible (w/ test) | Adverse Events | 745 (572 - 953) | 995 (765 - 1,310) | 1,830 (1,540 - 2,150) | 1,790 (1,550 - 2,070) | 2,060 (1,760 - 2,380) | 2,830 (2,430 - 3,270) | 3,120 (2,600 - 3,690) | 4,020 (3,370 - 4,750) |
| WPR | TPT for eligible (w/ test) | Serious Adverse Events | 319 (246 - 417) | 427 (321 - 560) | 785 (653 - 930) | 766 (652 - 900) | 884 (748 - 1,040) | 1,220 (1,010 - 1,440) | 1,340 (1,110 - 1,580) | 1,730 (1,430 - 2,070) |
| WPR | TPT for eligible (w/o test) | Adverse Events | 745 (572 - 953) | 1,900 (1,470 - 2,440) | 2,550 (2,160 - 3,000) | 2,360 (2,060 - 2,730) | 2,560 (2,210 - 2,930) | 3,330 (2,920 - 3,760) | 3,520 (3,060 - 4,070) | 4,430 (3,820 - 5,090) |
| WPR | TPT for eligible (w/o test) | Serious Adverse Events | 319 (246 - 417) | 816 (628 - 1,060) | 1,100 (917 - 1,290) | 1,010 (865 - 1,190) | 1,100 (935 - 1,280) | 1,430 (1,230 - 1,660) | 1,510 (1,280 - 1,750) | 1,900 (1,630 - 2,210) |

Table A18 Proportion and number of 177 countries in each cost-effectiveness category for including preventive treatment without testing up to each age group on top of HHCM with preventive treatment for those under 15 years.

ICER = incremental cost-effectiveness ratio; pcGDP = per capita gross domestic product.

| **ICER / pcGDP (%)** | **N, [15,25)** | **%, [15,25)** | **N, [25,35)** | **%, [25,35)** | **N, [35,45)** | **%, [35,45)** | **N, [45,55)** | **%, [45,55)** | **N, [55,65)** | **%, [55,65)** | **N, [65,Inf)** | **%, [65,Inf)** |
| --- | --- | --- | --- | --- | --- | --- | --- | --- | --- | --- | --- | --- |
| Cost-saving | 49 | 25.9 | 47 | 24.9 | 46 | 24.3 | 45 | 23.8 | 44 | 23.3 | 44 | 23.3 |
| <30% | 114 | 60.3 | 115 | 60.8 | 114 | 60.3 | 111 | 58.7 | 108 | 57.1 | 104 | 55 |
| 30-50% | 19 | 10.1 | 19 | 10.1 | 21 | 11.1 | 23 | 12.2 | 22 | 11.6 | 24 | 12.7 |
| 50-100% | 3 | 1.6 | 4 | 2.1 | 4 | 2.1 | 6 | 3.2 | 10 | 5.3 | 11 | 5.8 |
| 100-300% | 2 | 1.1 | 2 | 1.1 | 2 | 1.1 | 2 | 1.1 | 3 | 1.6 | 4 | 2.1 |
| 300-500% | 2 | 1.1 | 2 | 1.1 | 2 | 1.1 | 1 | 0.5 | 1 | 0.5 | 1 | 0.5 |
| >500% | 0 | 0 | 0 | 0 | 0 | 0 | 1 | 0.5 | 1 | 0.5 | 1 | 0.5 |

##### 3.0.2. Supplementary figures

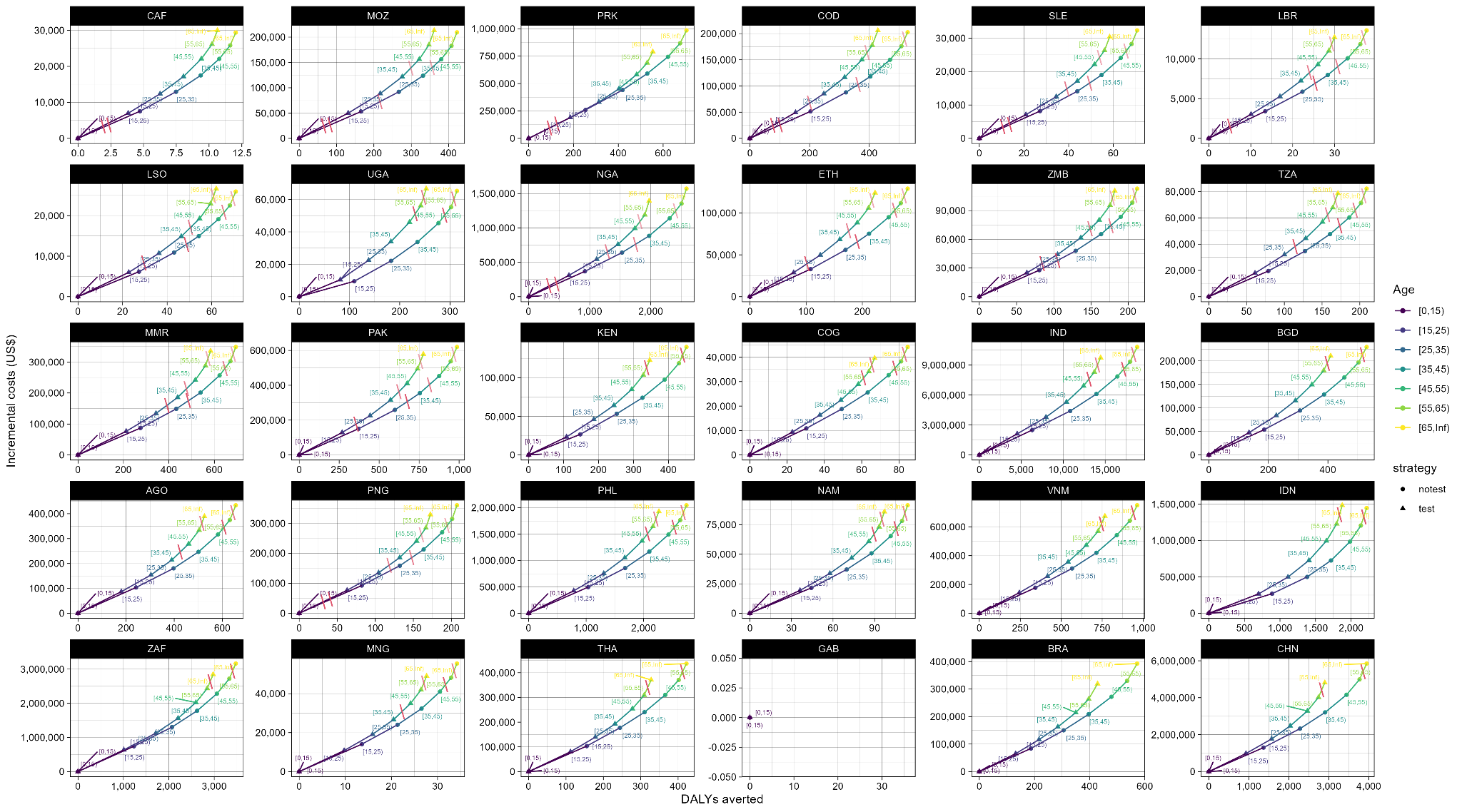

Figure A3 The incremental costs and benefits of providing tuberculosis preventive treatment to those eligible in each age group in addition to case-finding. Red lines indicate a per-capita gross domestic product (GDP) cost-effectiveness threshold; dashed red lines indicate a half-GDP cost-effectiveness threshold; dotted lines indicate a third-GDP cost-effectiveness threshold.
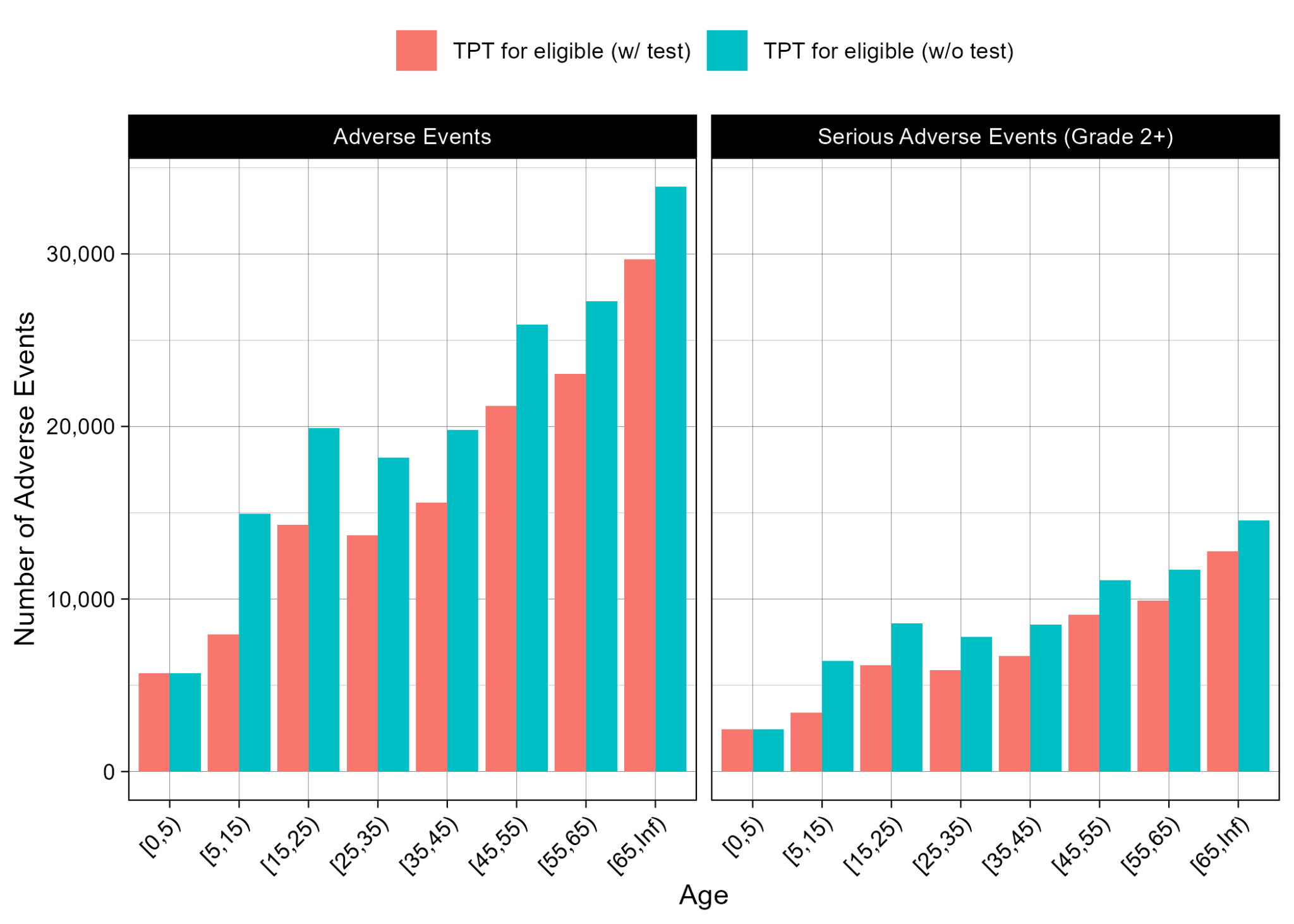

Figure A4 Estimated numbers of musculoskeletal adverse events and grade 2+ musculoskeletal adverse events associated with providing full tuberculosis preventive treatment coverage in each age group (see also Figure A5 and Table A17)

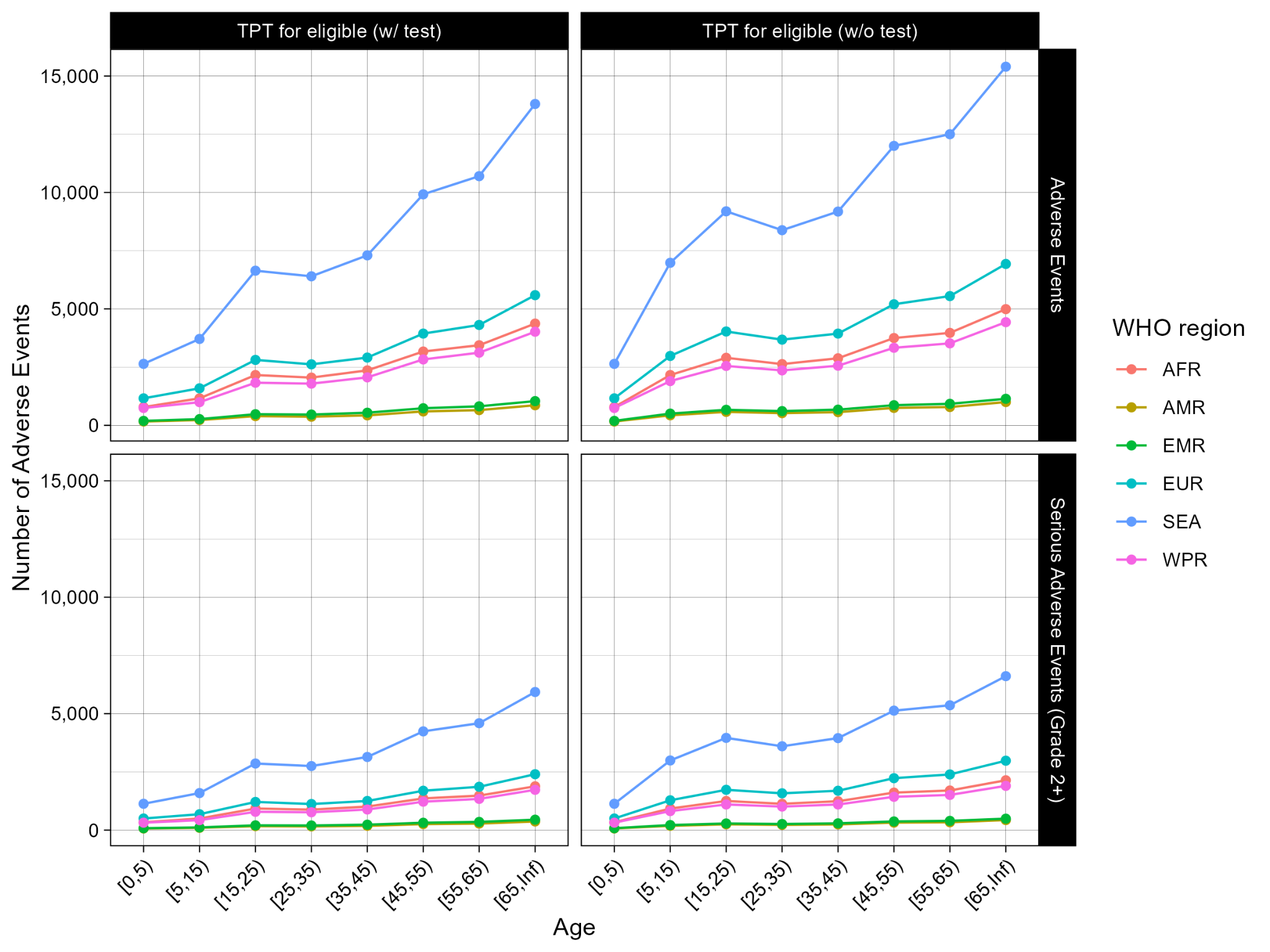

Figure A5 Estimated numbers of musculoskeletal adverse events and grade 2+ musculoskeletal adverse events associated with providing full tuberculosis preventive treatment coverage in each age group by WHO region (see also Figure A4 and Table A17)

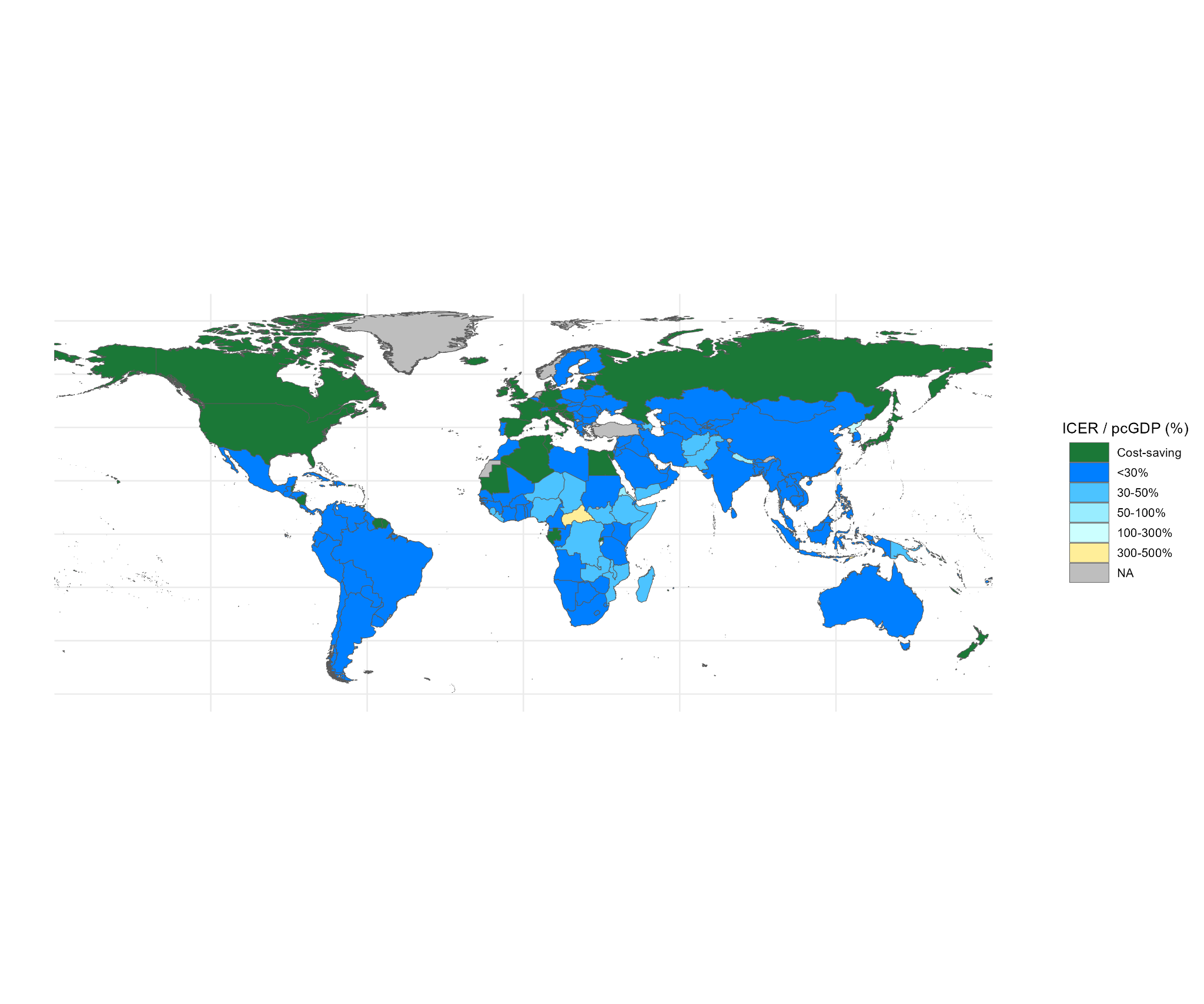

Figure A6 Incremental cost-effectiveness of adding tuberculosis preventive treatment without infection testing for contacts aged **under 25 years** onto multidrug-resistant/rifampicin-resistant tuberculosis household contact management with preventive treatment for those under 15 years. Grey denotes missing data.

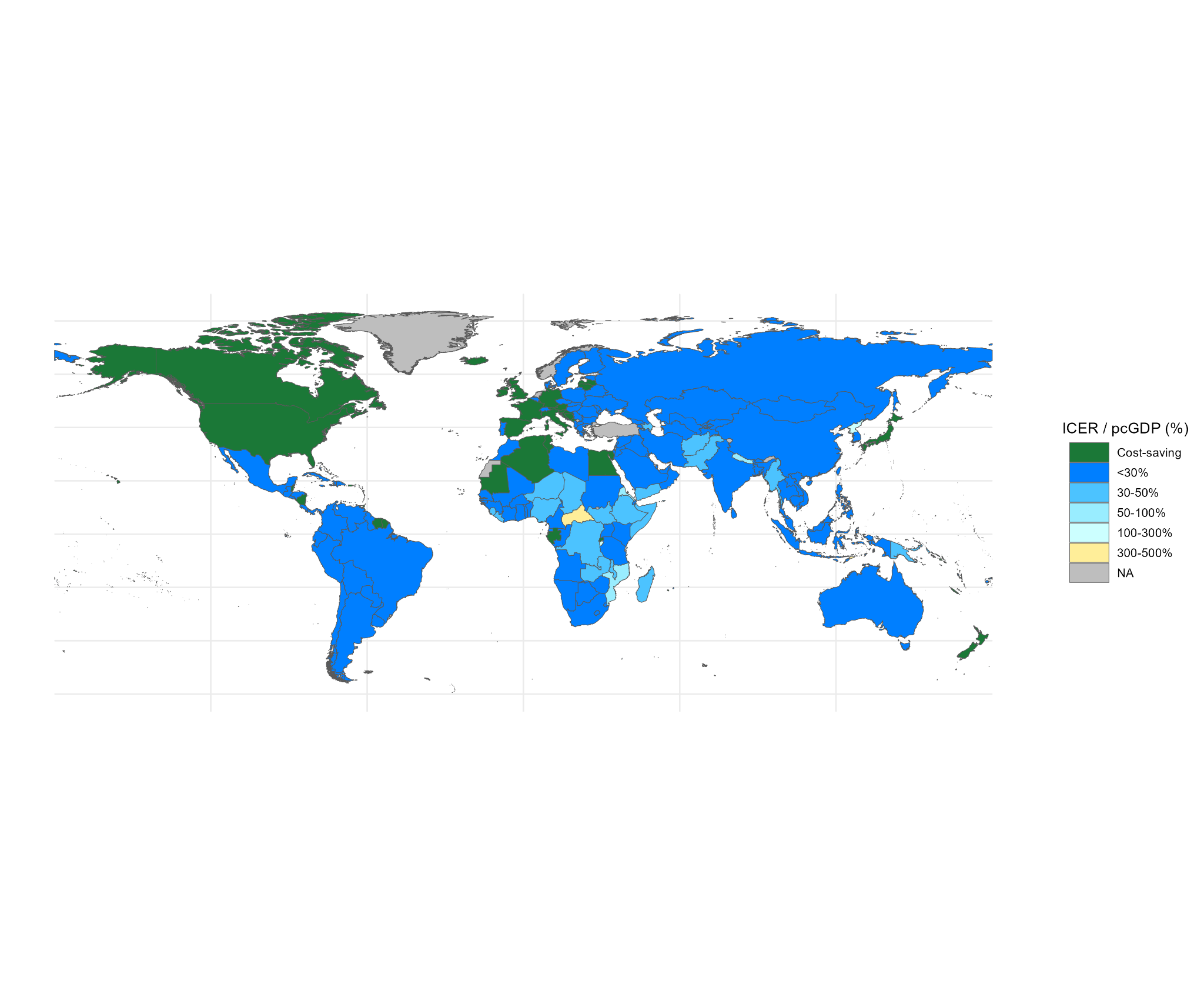

Figure A7 Incremental cost-effectiveness of adding tuberculosis preventive treatment without infection testing for contacts aged **under 35 years** onto multidrug-resistant/rifampicin-resistant tuberculosis household contact management with preventive treatment for those under 15 years. Grey denotes missing data.

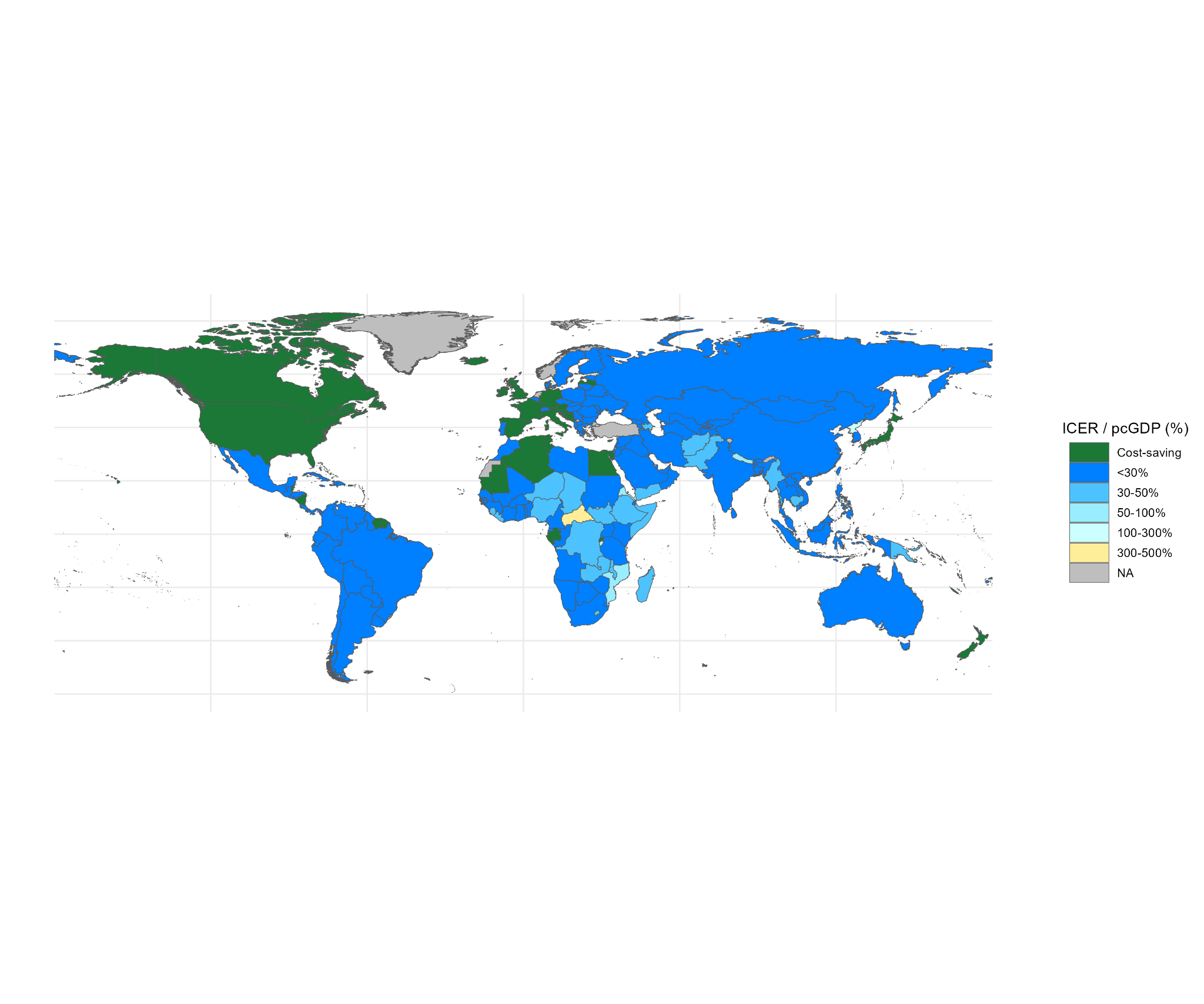

Figure A8 Incremental cost-effectiveness of adding tuberculosis preventive treatment without infection testing for contacts aged **under 45 years** onto multidrug-resistant/rifampicin-resistant tuberculosis household contact management with preventive treatment for those under 15 years. Grey denotes missing data. **Note:** this is the same as Figure 5 in the main article and is included here for ease of comparison.

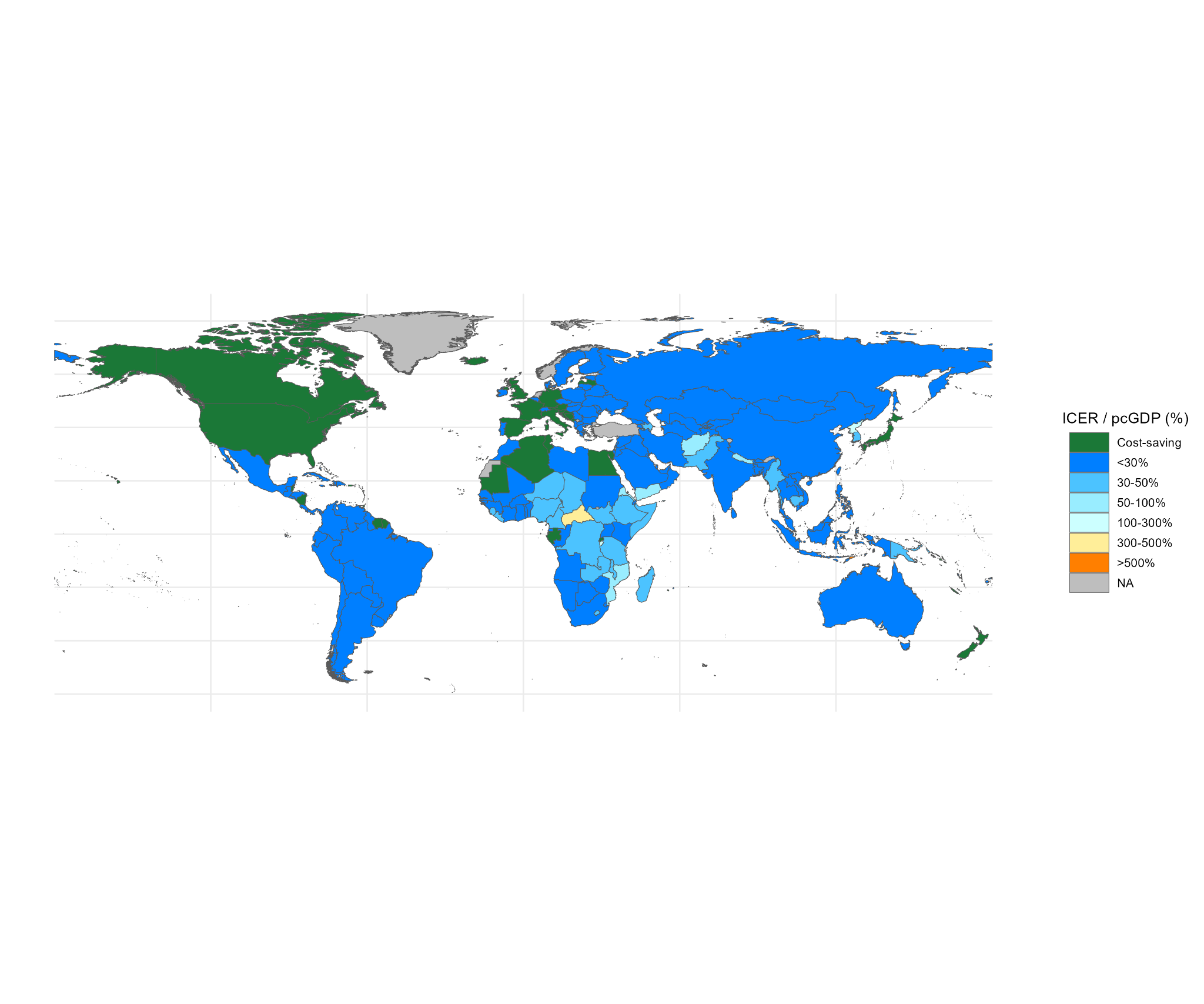

Figure A9 Incremental cost-effectiveness of adding tuberculosis preventive treatment without infection testing for contacts aged **under 55 years** onto multidrug-resistant/rifampicin-resistant tuberculosis household contact management with preventive treatment for those under 15 years. Grey denotes missing data.

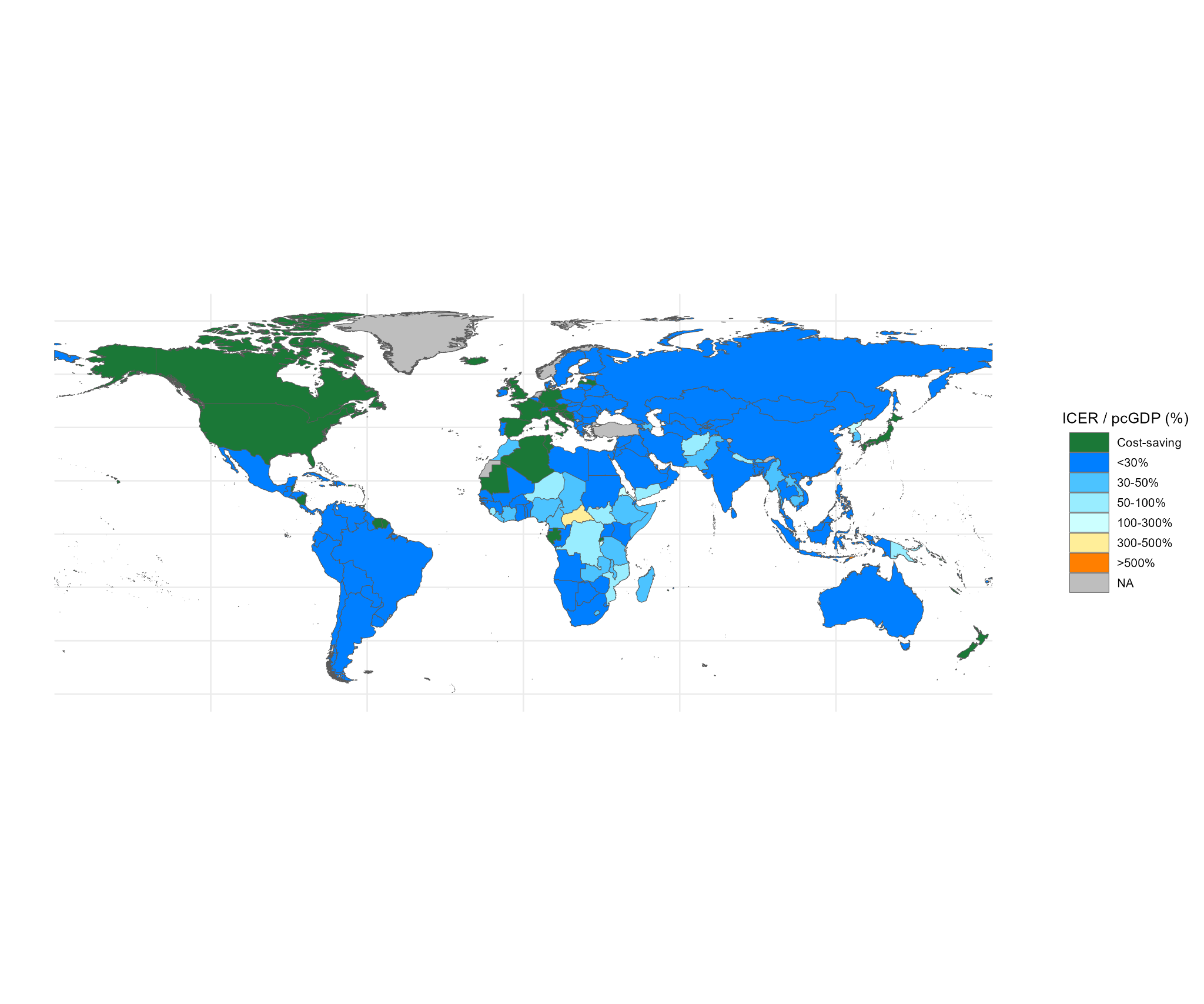

Figure A10 Incremental cost-effectiveness of adding tuberculosis preventive treatment without infection testing for contacts aged **under 65 years** onto multidrug-resistant/rifampicin-resistant tuberculosis household contact management with preventive treatment for those under 15 years. Grey denotes missing data.

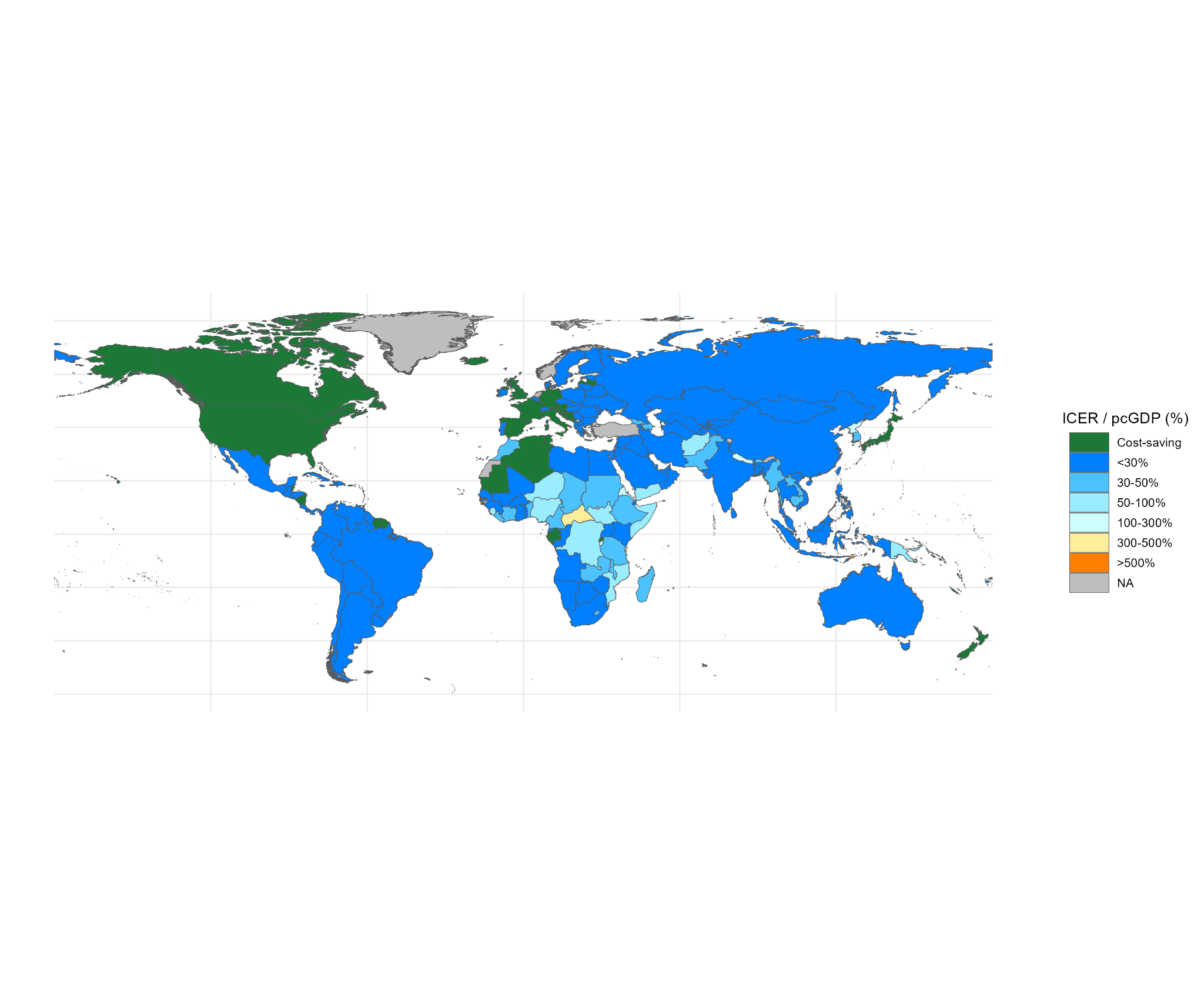

Figure A11 Incremental cost-effectiveness of adding tuberculosis preventive treatment without infection testing for contacts of **all ages** onto multidrug-resistant/rifampicin-resistant tuberculosis household contact management with preventive treatment for those under 15 years. Grey denotes missing data.
